# Linking latent trajectories of ageing-related atrophy, white matter hyperintensities, and cognitive ageing over four years: Insights into brain maintenance

**DOI:** 10.1101/2025.06.16.25329648

**Authors:** Inga Menze, Jose Bernal, Rogier A. Kievit, Renat Yakupov, Slawek Altenstein, Claudia Bartels, Katharina Buerger, Michaela Butryn, Peter Dechent, Michael Ewers, Klaus Fliessbach, Ingo Frommann, Maria Gemenetzi, Wenzel Glanz, Daria Gref, Julian Hellmann-Regen, Stefan Hetzer, Enise I. Incesoy, Daniel Janowitz, Ingo Kiliman, Luca Kleineidam, Marie Theres Kronmüller, Christoph Laske, Debora Melo van Lent, Falk Lüsebrink, Robert Perneczky, Oliver Peters, Lukas Preis, Josef Priller, Alfredo Ramirez, Boris-Stephan Rauchmann, Ayda Rostamzadeh, Sandra Roeske, Klaus Scheffler, Björn-Hendrik Schott, Anja Schneider, Sebastian Sodenkamp, Annika Spottke, Eike Jakob Spruth, Melina Stark, Stefan Teipel, Michael Wagner, Jens Wiltfang, Frank Jessen, Stefanie Schreiber, Emrah Düzel, Gabriel Ziegler

**Affiliations:** German Centre for Neurodegenerative Diseases (DZNE), Magdeburg, Leipziger Str. 44, 39120 Magdeburg, Germany; Institute of Cognitive Neurology and Dementia Research, Otto-von-Guericke University Magdeburg, Magdeburg, Leipziger Str. 44, 39120 Magdeburg, Germany; Centre for Clinical Brain Sciences, The University of Edinburgh, 49 Little France Crescent, EH16 4SB Edinburgh, UK; University of Edinburgh and UK DRI, Edinburgh, Edinburgh Bioquarter, 49 Little France Crescent, Edinburgh, EH16 4SB, UK; Donders Institute for Brain, Cognition and Behavior, Radboud University Medical Center, Trigon Building, Kapittelweg 29, Nijmegen, 6525 EN, the Netherlands; German Center for Neurodegenerative Diseases (DZNE), Berlin, Charitéplatz 1, 10117 Berlin, Germany; Department of Psychiatry and Psychotherapy, Charité, Charitéplatz 1, 10117 Berlin, Germany; Department of Psychiatry and Psychotherapy, University Medical Center Goettingen, University of Goettingen, Von-Siebold-Str. 5, 37075 Goettingen; German Center for Neurodegenerative Diseases (DZNE, Munich), Feodor-Lynen-Strasse 17, 81377 Munich, Germany; Institute for Stroke and Dementia Research (ISD), University Hospital, LMU Munich, Feodor-Lynen-Strasse 17, 81377 Munich, Germany; MR-Research in Neurosciences, Department of Cognitive Neurology, University Medical Center Goettingen, Robert-Koch-Straße 40, 37075 Goettingen, Germany; German Center for Neurodegenerative Diseases (DZNE), Bonn, Venusberg-Campus 1, 53127 Bonn, Germany; Department of Old Age Psychiatry and Cognitive Disorders, University Hospital Bonn, Venusberg-Campus 1, 53127 Bonn, Germany; Charité Universitätsmedizin Berlin, Department of Psychiatry and Neurosciences, Hindenburgdamm 30, 12203 Berlin; Charité Universitätsmedizin Berlin, ECRC Experimental and Clinical Research Center, Lindenberger Weg 80, 13125 Berlin, Germany; Berlin Center for Advanced Neuroimaging, Charité – Universitätsmedizin Berlin, Charitéplatz 1, 10117 Berlin, Germany; Vivantes Klinikum Am Urban, Department of Psychiatry, Psychotherapy, and Psychosomatics, Vivantes Urban Hospital, Dieffenbachstraße 1, 10967 Berlin, Germany; German Center for Neurodegenerative Diseases (DZNE), Rostock, Gehlsheimer Str. 20, 18147 Rostock, Germany; Department of Psychosomatic Medicine, Rostock University Medical Center, Gehlsheimer Str. 20, 18147 Rostock; German Center for Neurodegenerative Diseases (DZNE), Tübingen, Otfried-Müller-Straße 23, 72076 Tübingen, Germany; Section for Dementia Research, Hertie Institute for Clinical Brain Research and Department of Psychiatry and Psychotherapy, University of Tübingen, Osianderstraße 24, 72076 Tübingen, Germany; Glenn Biggs Institute for Alzheimer’s and Neurodegenerative Diseases, UT Health San Antonio, 8300 Floyd Curl Drive, San Antonio Texas 78229, USA; Department of Psychiatry and Psychotherapy, University Hospital, LMU Munich, Nussbaumstrasse 7, 80336 Munich, Germany; Munich Cluster for Systems Neurology (SyNergy) Munich, Feodor-Lynen-Strasse 17, 81377 Munich, Germany; Ageing Epidemiology Research Unit (AGE), School of Public Health, Imperial College London, Charing Cross Hospital, St Dunstan’s Road, London W6 8RP, UK; Department of Psychiatry and Psychotherapy, School of Medicine and Health, Technical University of Munich, and German Center for Mental Health (DZPG), Ismaninger Straße 22, 81675 Munich, Germany; Excellence Cluster on Cellular Stress Responses in Aging-Associated Diseases (CECAD), University of Cologne, Joseph-Stelzmann-Strasse 26, 50931 Köln, Germany; Division of Neurogenetics and Molecular Psychiatry, Department of Psychiatry and Psychotherapy, Faculty of Medicine and University Hospital Cologne, University of Cologne, Kerpener Str. 62, 50931 Cologne, Germany; Department of Psychiatry & Glenn Biggs Institute for Alzheimer’s and Neurodegenerative Diseases, San Antonio, 8300 Floyd Curl Drive, San Antonio Texas 78229, USA; Sheffield Institute for Translational Neuroscience (SITraN), University of Sheffield, Western Bank, Sheffield, S10 2TN, UK; Department of Neuroradiology, University Hospital LMU, Marchioninistr. 15, 81377 Munich, Germany; Department of Psychiatry, University of Cologne, Medical Faculty, Kerpener Strasse 62, 50924 Cologne, Germany; Department for Biomedical Magnetic Resonance, University of Tübingen, Otfried-Müller-Straße 51, 72076 Tübingen, Germany; German Center for Neurodegenerative Diseases (DZNE), Goettingen, Von-Siebold-Str. 3a, 37075 Goettingen, Germany; Leibniz Institute for Neurobiology, Brenneckestr. 6, 39118, Magdeburg, Germany; Department of Psychiatry and Psychotherapy, University of Tübingen, Osianderstraße 24, 72076 Tübingen, Germany; Department of Neurology, University of Bonn, Venusberg-Campus 1, 53127 Bonn, Germany; Neurosciences and Signaling Group, Institute of Biomedicine (iBiMED), Department of Medical Sciences, University of Aveiro, Campus Universitario de Santiago, Agra do Crasto – Edificio 30, 3810-193 Aveiro, Portugal; Department of Neurology, University Hospital Magdeburg, Magdeburg, Leipziger Str. 44, 39120 Magdeburg, Germany

**Keywords:** Brain maintenance, white matter hyperintensities, ageing-related atrophy, cognitive decline, modifiable lifestyle factors, longitudinal analysis, structural equation modelling, latent growth curve modelling, multicentre study

## Abstract

**Introduction:** We studied brain maintenance, examining the co-evolution of ageing-related atrophy, white matter hyperintensities (WMH), and cognition, alongside domain-specific and domain-general contributions of lifestyle and personality.

**Methods:** In 543 cognitively unimpaired DELCODE participants, we modelled four-year interrelations between medial temporal lobe to ventricle ratio (MTLV-ratio), WMH, and PACC5 performance using latent growth curve modelling. We quantified unique contributions of brain changes to cognitive change and derived a domain-general brain maintenance index. Associations with lifestyle and personality were examined post-hoc.

**Results:** Steeper MTLV-ratio decline related to baseline WMH and its progression. Brain-domain changes independently contributed to cognitive changes. Neuroticism, depressive symptoms, and low cognitive engagement related to unfavourable domain-specific trajectories and brain maintenance index.

**Discussion:** Dynamics of WMH and ageing-related atrophy on cognitive ageing highlight their relevance for brain maintenance. Our results suggest that maintaining cerebrovascular and mental health alongside cognitive engagement could promote brain maintenance, delay cognitive decline and dementia.

## 1 Introduction

Ageing is accompanied by marked structural and functional brain changes, including altered connectivity, atrophy, and network reorganisation ^1,2^. Ageing-related atrophy, especially in prefrontal regions and the medial temporal lobe (MTL) ^3–5^, is a hallmark of normal ageing and often coincides with ventricular enlargement, reflecting parenchymal tissue loss ^4,6^. Cerebrovascular abnormalities, commonly attributed to cerebral small vessel disease (CSVD), also become increasingly prevalent with age ^3,7,8^. Among them, white matter hyperintensities (WMH) have received particular attention due to their high prevalence in older adults, links to cardiovascular risk, and robust association with cognitive decline ^9,10^.

Together, structural and cerebrovascular brain changes may contribute to cognitive decline, although their relative impact and expression vary substantially across individuals ^11^. Substantial heterogeneity has been observed in cerebrovascular abnormalities ^7,12,13^, ageing-related structural brain changes ^11,12^, and cognitive trajectories ^11,12,14^. Elucidating the factors underlying this variability and the dynamic interplay among these domains is critical for advancing mechanistic models of neurocognitive ageing and informing strategies to prevent cognitive decline in older adults.

The concept of brain maintenance offers a theoretical framework for investigating individual differences in neurocognitive ageing ^15,16^. Brain maintenance refers to the relative preservation of brain structure and function by attenuating ageing- or pathology-related changes. The framework seeks to identify genetic, sociodemographic, and lifestyle factors that promote such preservation ^16^, thus supporting cognitive abilities throughout ageing via joint neurocognitive changes ^15,16^. Multiple modifiable lifestyle factors have been linked to cognitive and brain health outcomes ^17^, including cognitive, social, and physical activity ^18,19^, cardiovascular risk factors ^17^, dietary patterns ^20^, as well as psychological risk factors like depression ^17,21,22^. While these partially modifiable lifestyle factors have primarily been linked to specific neuroimaging-derived brain outcomes, it is also plausible that maintaining integrity in one domain—such as cerebrovascular health—may support the preservation of another, e.g. MTL structure. Moreover, such distinct processes may also co-evolve over time in interdependent trajectories. In this context, our study focusses on modelling the intricate relationships between cerebrovascular abnormalities and age-related brain atrophy, and their specific contribution to cognitive decline in ageing.

Although cerebrovascular alterations and ageing-related brain atrophy often co-occur in ageing, the nature of their interrelationship remains debated ^23,24^. While some studies report no significant associations between WMH and brain atrophy ^25^, others attribute their co-occurrence to shared risk factors, such as Alzheimer’s disease (AD) biomarkers ^26,27^. Conversely, bidirectional models propose that WMH and atrophy exacerbate one another over time ^13,28,29^. WMH may also directly contribute to brain atrophy ^30,31^, particularly in MTL structures ^24,30^, highlighting the prevention of cerebrovascular abnormalities as a potential target for preserving vulnerable brain structures.

The unresolved complexities of their dynamics hinder understanding of the unique and synergistic effects of WMH and ageing-related atrophy on cognitive decline, especially in longitudinal contexts. Some prospective studies in older adults have demonstrated independent ^25^ or interactive effects ^32,33^ of baseline markers of ageing-related atrophy and cerebrovascular abnormalities on cognitive trajectories, with outcomes depending on age group and cognitive domain. Studies examining simultaneous changes in these neurocognitive domains yielded ambiguous results. Some suggest interactive effects of hippocampal atrophy and WMH progression on episodic and working memory changes over nine years ^34^, while others associate episodic memory changes over 15 years primarily with four-year hippocampal atrophy ^12^. In a study examining simultaneous change over three years, changes in WMH, total brain-, grey- and white matter volume, and cognition were significantly interrelated ^31^.

In this study, we investigated the longitudinal interrelations among the three domains age-related brain atrophy, cerebrovascular abnormalities, and cognitive performance over four years in a large sample of cognitively unimpaired older adults assessed annually. Leveraging multivariate latent growth curve modelling within the brain maintenance framework, we examined shared changes across these three neurocognitive domains, while accounting for common risk factors. We also assessed the unique contributions of brain domain changes to cognitive domain changes. Finally, we explored both domain-specific and domain-general contributors to brain maintenance by evaluating how modifiable lifestyle factors and personality traits relate to individual differences in baseline levels and longitudinal change across these three neurocognitive domains.

## 2 Methods

### 2.1 Study design and participants

This study focussed on baseline and annual follow-up data up to 48 months of 543 cognitively unimpaired individuals from DELCODE (DZNE Longitudinal Cognitive Impairment and Dementia Study ^35^)—an observational multicentre study from the German Centre for Neurodegenerative Diseases (DZNE). We restricted our analysis to individuals who successfully completed at least two visits within the period of 2014 to 2023.

Subjects in DELCODE were ≥ 60 years old and considered cognitively unimpaired, if they performed above -1.5 SD of normal performance on all subtests of the Consortium to Establish a Registry for Alzheimer’s Disease (CERAD) plus test battery adjusting for age, sex, and education. Additional exclusion and inclusion criteria have been previously described in more detail ^35^.

All participants gave written informed consent in accordance with the Declaration of Helsinki prior joining the study. DELCODE is retrospectively registered at the German Clinical Trials Register (DRKS00007966, 04/05/2015).

### 2.2 Brain imaging data and domains of brain structure

MRI acquisition took place at nine DZNE neuroimaging sites using 3T Siemens MR scanners. We used structural scans in terms of T1w MPRAGE (full head coverage; 3D acquisition, GRAPPA factor 2, 1 mm^3^ isotropic, 256 × 256 px, 192 sagittal slices, TR/TE/TI 2500/4.33/1100 ms, FA 7°) and T2w FLAIR (full head coverage; 1 mm^3^ isotropic, 256 × 256 px, 192 sagittal slices, TR/TE/TI 5000/394/1800 ms). The DZNE imaging network oversaw operating procedures and quality assurance and assessment (iNET, Magdeburg) ^35^.

#### 2.2.1 Quantification of ageing-related medial temporal lobe atrophy

We used Freesurfer’s longitudinal pipeline for segmentation in T1w MPRAGE images. We specifically focused on the aggregated volumes of the hippocampus, entorhinal cortex, parahippocampal cortex, and amygdala, which are broadly considered key structures of the MTL, as well as volumes of the inferior lateral ventricles. As a measure of ageing-related brain atrophy and MTL-integrity, we computed the MTL-to-ventricle ratio (MTLV-ratio):

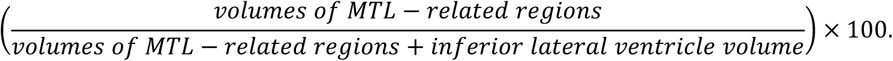

Similar measures, such as the previously proposed hippocampal-to-ventricle ratio have been shown to be more sensitive to ageing and cognitive decline than absolute volume measures of brain tissue ^36,37^.

We estimated segmentation-based total intracranial volume (TICV) from baseline, which relies on Freesurfer’s samseg-based structure segmentation ^38^, including CSF and other intra-cranial non-brain structures (https://surfer.nmr.mgh.harvard.edu/fswiki/sbTIV).

#### 2.2.2 White matter hyperintensities segmentation

To characterize individual cerebrovascular abnormalities, we used LST-AI ^39^, along with T1w MPRAGE and T2w FLAIR imaging, to quantify the individual total WMH volume of all participants and time points.

### 2.3 Cognitive domain

We assessed cognitive performance annually using the preclinical Alzheimer’s cognitive composite score (PACC5) ^40^, a measure for early cognitive change specific for AD. The PACC5 comprises measures of processing speed, global cognition, and particularly memory—cognitive domains that are particularly vulnerable to AD pathology but also show age-related decline in cognitively healthy individuals. The PACC5 is the averaged z-standardised performance on the Mini Mental State Examination (MMSE), Wechsler Memory Scale Revised (WMS-R) logical memory delayed recall, Symbol-Digit-Modalities-Test (SDMT), free and total recall of the Free and Cued Selective Reminding Test (FCSRT), and semantic fluency. Individual-level z-scores were derived from baseline mean and SD of the cognitively unimpaired individuals.

### 2.4 Modifiable lifestyle factors and personality traits

To examine contributors to trajectories of each neurocognitive domain (domain-specific) or across them (domain-general) in the context of brain maintenance, we assessed multiple potentially modifiable lifestyle factors via self-report questionnaires, including cardiovascular risk, late-life depressive symptoms, Mediterranean diet (MeDi), physical activity, sleep quality, social network, and lifetime experiences. Additionally, we assessed personality traits. Some of these factors were collected once during the baseline visit and some on a continuous basis during each annual visit (overview in **Supplementary Table 1**). As some individuals did not have measurements for specific variables of interest at baseline but provided ratings at later time points (**Supplementary** Figure 1), we opted to compute the mean of all available measurements to capture the average manifestation of the lifestyle factor throughout the study period.

### 2.5 Statistical analysis

The goals of our model-based analysis of joint neurocognitive changes to study brain maintenance were two-fold: First, we established linked changes across the three neuro-cognitive domains and explored unique contributions of changes in brain structure domains to cognitive ageing. Second, we sought to identify modifiable lifestyle factors and personality traits that contributed domain-specific or domain-general to the preservation of these linked neurocognitive domains in ageing.

#### 2.5.1 Data transformation

To account for undesired effects of potential skewness, we log10-transformed WMH volumes, Box-Cox transformed MTLV-ratio, and Yeo-Johnson transformed PACC5. All variables were z-scored (pooled across time points) before entering the model.

#### 2.5.2 Latent Growth Curve Modelling

For multi-domain trajectory modelling and maintenance analysis we leveraged latent growth curve modelling (LGCM) ^41^, a flexible and powerful class of structural equation models (SEM). This approach allows for the estimation of both latent baseline levels (intercepts) and longitudinal change (slopes) in constructs of interest, while accounting for covariate effects. In the context of this study, we use the term domain to refer to the latent constructs representing WMH, MTLV-ratio, and cognitive performance. Specifically, we employed trivariate LGCM to jointly analyse longitudinal trajectories and interrelations between WMH, MTLV-ratio, and cognitive performance. Specifically, the LGCM allowed us to examine the following:

1. How are the baseline levels of WMH, MTLV-ratio, and cognitive performance associated with one another (covariance between latent intercepts)?
2. How do WMH, MTLV-ratio, and cognitive performance change over the course of four years, and is there interindividual variability in latent change (slopes)?
3. Are the latent intercepts and latent slopes of each domain associated to covariates?
4. Do baseline levels in one domain affect changes in the other two (covariance between latent intercepts and latent slopes across domains). We were particularly interested if baseline levels of WMH affected MTLV-ratio decline rates and cognitive decline, as well as if baseline levels of MTLV-ratio related to cognitive decline.
5. Are changes in one domain associated to changes in another (covariance between latent slopes)?

All latent intercepts and latent slopes were adjusted for effects of age, sex, years of education. Moreover, latent intercepts and latent slopes of WMH and MTLV-ratio were corrected for TICV.

We modelled linear latent slopes resulting in rates of change per year. Models were fitted using robust maximum likelihood estimator (MLR) and missing data were handled using Full Information Maximum Likelihood Estimation (FIML). We ascertained the assumption of data missing at random via Little’s missing completely at random test (χ2(1466) = 1544.30, *p* = 0.076).

Prior to model fitting, we identified and removed outliers that were above Q3 + 1.5×IQR or below Q1 - 1.5×IQR of the median for WMH, MTLV-ratio, and PACC5 performance, respectively, pooling data across time points.

The overall model fit was evaluated by the χ² test, Comparative Fit Index (CFI), root mean square error of approximation (RMSEA), and standardized root mean square residuals (SRMR). Good model fit was defined as CFI ≥ 0.97, RMSEA ≤ 0.05 , SRMR ≤ 0.05. We reported estimates from the fully standardized solution to present comparable, unit-independent effect estimates. We report full model information including the raw estimates in supplements. The threshold for *p*-values was set to *p* ≤ 0.05.

To facilitate the assessment of the number of individuals progressing or regressing in either domain, we extracted regression-based factor score estimates of latent slopes for each individual and report the percentages of individuals with negative or positive factor scores.

##### 2.5.2.1 Supplementary analysis I: Assessing contribution of shared risk factors

Relationships between the neurocognitive domains of interest were proposed to possibly stem from shared factors, such as cardiovascular risks ^23^. Moreover, in the context of Alzheimer’s disease (AD), brain atrophy and WMH progression might be entangled due to the accumulation of AD biomarkers ^26,27^, while alternative hypotheses claim they are entirely distinct and independent epiphenomena ^42,43^. In supplementary analyses, we therefore tested a LGCM which additionally corrected for (a) vascular risk on latent intercepts and latent slopes of WMH and MTLV-ratio, and (b) APOE-ε4 carriership and plasma Aβ42/40 on all latent intercepts and slopes ^44^. We examined whether controlling for these common underlying risk factors would alter the covariant structure between neurocognitive domains.

##### 2.5.2.2 Supplementary analysis II: Differences between converting and cognitively stable individuals

To validate our multi-domain model, we examined in a supplementary analyses whether latent factor scores in the three neurocognitive domains of interest differed between individuals who stayed cognitively stable within the course of the study (n=413) and those who converted to mild cognitive impairment (MCI) or dementia (n=93 [n_converted to MCI_=86, n_converted to MCI and subsequent dementia_=7]) Conversion status in all individuals was assessed up to 5 years and 4 months after inclusion in the study (mean follow up time: 5.41 years; n = 386; 37 individuals exceeded this range). Information on clinical progression to MCI or dementia was assessed for cognitively unimpaired individuals up until April 2023 ^45^.

##### 2.5.2.3 Unique contributions of WMH and MTLV-ratio rates of change to cognitive ageing

We aimed to examine the independent contribution of changes in WMH and MTLV-ratio to cognitive change, as the covariance of two latent variables LGCM does not account for the effect of the other latent effects. To facilitate interpretation, we divided our analysis into two parts. First, we estimated latent intercepts and latent slopes (see 2.5.2) and extracted regression-based factor score estimates for each individual. Second, we used multiple linear regression to examine the effect of WMH and MTLV-ratio latent slopes on the latent slopes of PACC5 performance, adjusting also for all latent intercepts. We identified and excluded potential outliers (*n* = 38) based on studentized residuals (cut-off ±3) and influential data points based on Cook’s distance (4/n; ^46^). We tested a model with only additive effects against a model including an interaction effect of the latent slopes of WMH and MTLV-ratio to examine possible synergistic effects of both pathological processes on cognitive changes. For model comparison, we used *F*-test and *ΔAIC* (*ΔAIC* ≤ 2 denoting no substantial difference between models).

#### 2.5.3 Domain-specific and domain-general contributors to neurocognitive changes

Using the extracted factor scores, we examined domain-specific relationships with modifiable lifestyle factors and personality traits via FDR-corrected Spearman’s correlation that accounted for sex, age, years of education, and TICV. We also report unaccounted correlations in supplements. In order to give an estimate of how much these contributing factors may explain in the variability of factor scores, we leveraged multiple linear regression. We identified and excluded potential outliers based on studentized residuals (cut-off ±3) and influential data points based on Cook’s distance (4/n; ^46^) for each model before fitting.

Based on research suggestions for brain maintenance ^16^, we additionally quantified maintenance more explicitly using the predicted values from the multiple regression model (see 2.5.3.2), which are thought to reflect the brain-structure–related component of cognitive ageing. The index describes individual differences of cognitive changes which are shared with structural brain changes. Higher individual predicted values hence indicate preserved cognitive functioning related to preserved brain integrity, consistent with more successful brain maintenance. We then correlated this brain maintenance index with modifiable lifestyle factors and personality traits to identify domain-general contributors to brain maintenance.

##### 2.5.3.1 Supplementary analysis III: Differences between individuals with high and low education

As lifestyle could counterbalance the risk for cognitive decline and brain integrity in face of low education or socioeconomic status ^47^, we explored whether associations between lifestyle factors and personality traits and each neurocognitive domain differed between individuals with high and low education—a key factor for preserving cognition and lower dementia risk ^17,48^. We divided the sample into individuals with lower vs. higher education based on a median split (*median_years of education_* = 14; lower education: *n* = 273, *mean_years of education_* = 12.3 ± 1.29; higher education: *n* = 270, *mean_years of education_* = 17.4 ± 1.60). To ensure that education-related variability in latent slopes of WMH, MTLV-ratio, and PACC5 performance was retained, we retrieved regression-based factor scores from a trivariate LGCM which did not include years of education. We then computed the aforementioned FDR-corrected Spearman’s correlations between lifestyle and domain-specific latent slopes in lower and higher education group separately, accounting for sex, age, and TICV.

#### 2.5.4 Software

We carried out all analyses in *R* (v4.2.3) using RStudio (v1.3.1073). We modelled trivariate LGCM using *lavaan* (v0.6-16). We used Mann-Whitney-U tests in *rstatix* and partial correlation in *ppcor*. We created figures using *ggplot2* (v3.4.2) and *semPlot* (v1.1.6).

## 3 Results

### 3.1 Descriptive statistics and sample characteristics

Among the 722 cognitively unimpaired DELCODE participants, 543 attended a minimum of two annual visits (52.85% female; mean age 69.99 ± 5.87 years; mean years of education: 14.82 ± 2.92 years; 27.88 % APOE4 carriers). The average number of visits attended per participant was approximately four (3.79, 95%-CI [3.70, 3.88]). Descriptive statistics of personality traits and modifiable lifestyle factors are detailed in **Supplementary Table 2**.

### 3.2 Longitudinal interrelations across neurocognitive domains of ageing-related atrophy, WMH, and cognition

In order to characterize latent-level interrelated changes in the three neurocognitive domains, we specified a trivariate LGCM model showing a good model fit (*χ²*(151) = 213.56, *p* = 0.001; *CFI* = 0.995; *RMSEA* = 0.028; *SRMR* = 0.018; **Figure 1A**; see **Supplementary Table 3** for detailed model information). Models additionally including the assumed shared risk factors cardiovascular risk, and APOE-ε4 and Plasma Aβ42/40 are presented in the supplements (**Supplementary** Figure 2; **Supplementary Table 4&5**). Since accounting for these additional covariates neither significantly improved model fit nor altered the interpretation of the results (**Supplementary Table 4&5**), we present and interpret the results of the parsimonious model, including only the covariates age, sex, years of education, and TICV.

**Figure 1.**
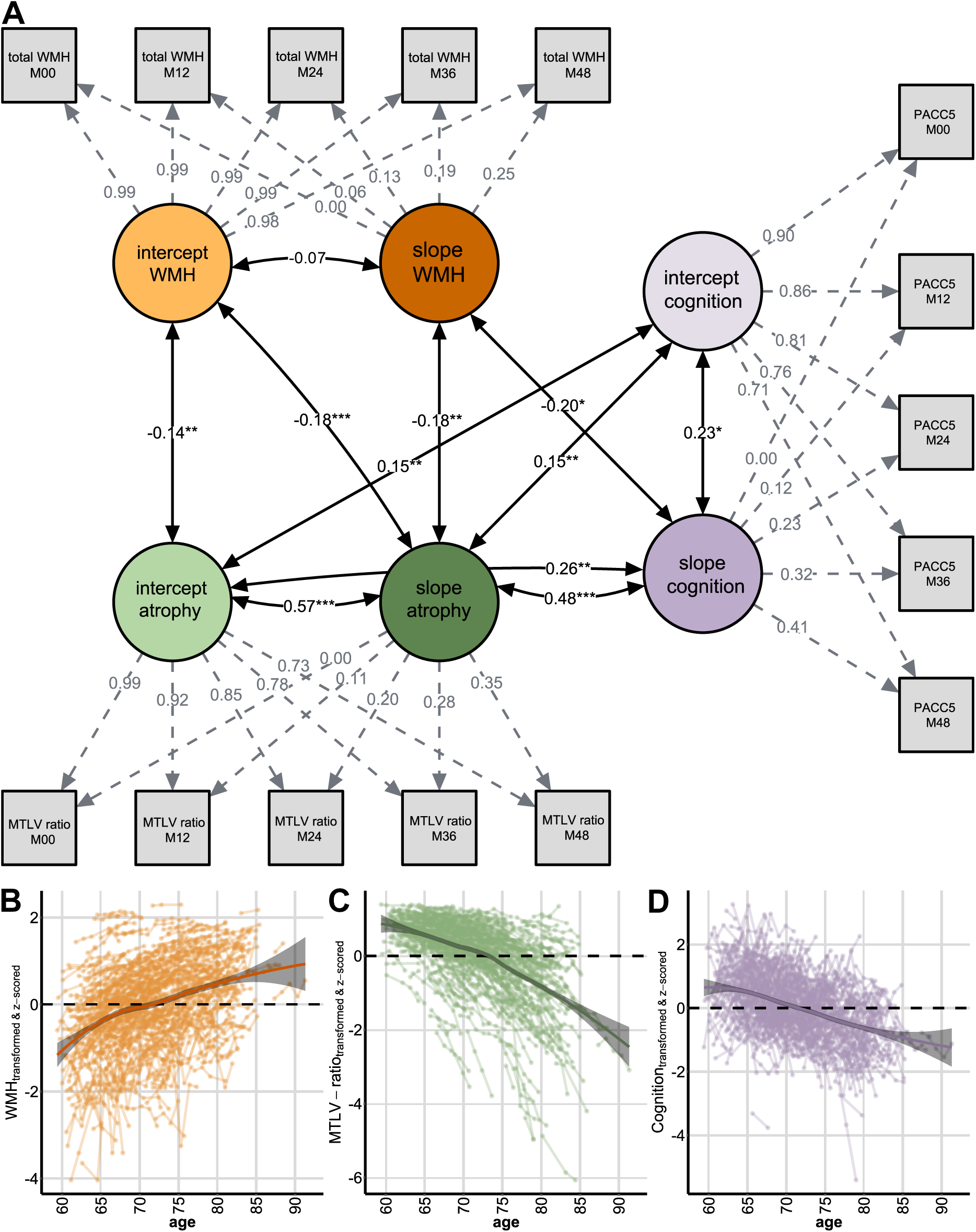
Illustration of the trivariate latent growth curve model linking changes in three neurocognitive domains. **(A)** We examined the interrelationship between three processes: cerebrovascular abnormalities, atrophy, and cognition, operationalized here as white matter hyperintensities (WMH), medial temporal lobe to ventricle ratio (MTLV-ratio), and PACC5 performance, respectively. We adjusted the model for age, sex, years of education, and in the case of WMH and MTLV-ratio for total intracranial volume (TICV). For readability we here do not show regressions of the covariates on latent intercepts and latent slopes (for information see **Supplementary Table 2**). Model shows the standardized coefficients. Fixed paths are depicted via dotted lines. For readability, only significant or trend-wise associations are depicted. * p < 0.05, ** p < 0.01, *** p < 0.001. **(B)** Individual trajectories for WMH over ageing. Total WMH volumes were log10-transformed and z-scored. **(C)** Individual trajectories for MTLV-ratio over ageing. MTLV-ratio was Box-Cox-transformed and z-scored. **(D)** Individual trajectories for PACC5 performance over ageing. PACC5 performance was Yeo-Johnson transformed and z-scored.

#### 3.2.1 Neurocognitive changes over time

On average, WMH volumes increased, MTLV ratios decreased, and PACC5 performance improved throughout the study period (intercept of WMH slope: *B* = 1.084, *Z* = 16.77, *p* < 0.001, **Figure 1B**; intercept of MTVL-ratio slope*: B* = -1.269, *Z* = -27.212*, p* < 0.001, **Figure 1C**; intercept of cognition slope*: B* = 0.314, *Z* = 4.131, *p* < 0.001, **Figure 1D**). Specifically, 89.69% of participants exhibited WMH progression, 93.55% an MTLV-ratio decline, and 66.48% positive PACC5 changes. PACC5 performance increases over follow-ups are likely to reflect a mixture of practice effects and ageing-related cognitive changes, and we refer to them as PACC5 performance changes from hereon. Importantly, both the starting points and the rate at which the aforementioned neurocognitive domains changed varied significantly across individuals (**Table 1**), suggesting that relative differences may provide meaningful insight into individual trajectories of neurocognitive ageing. **Supplementary** Figure 3 shows the differences in rates of change between cognitively stable and converted individuals.

**Table 1.**
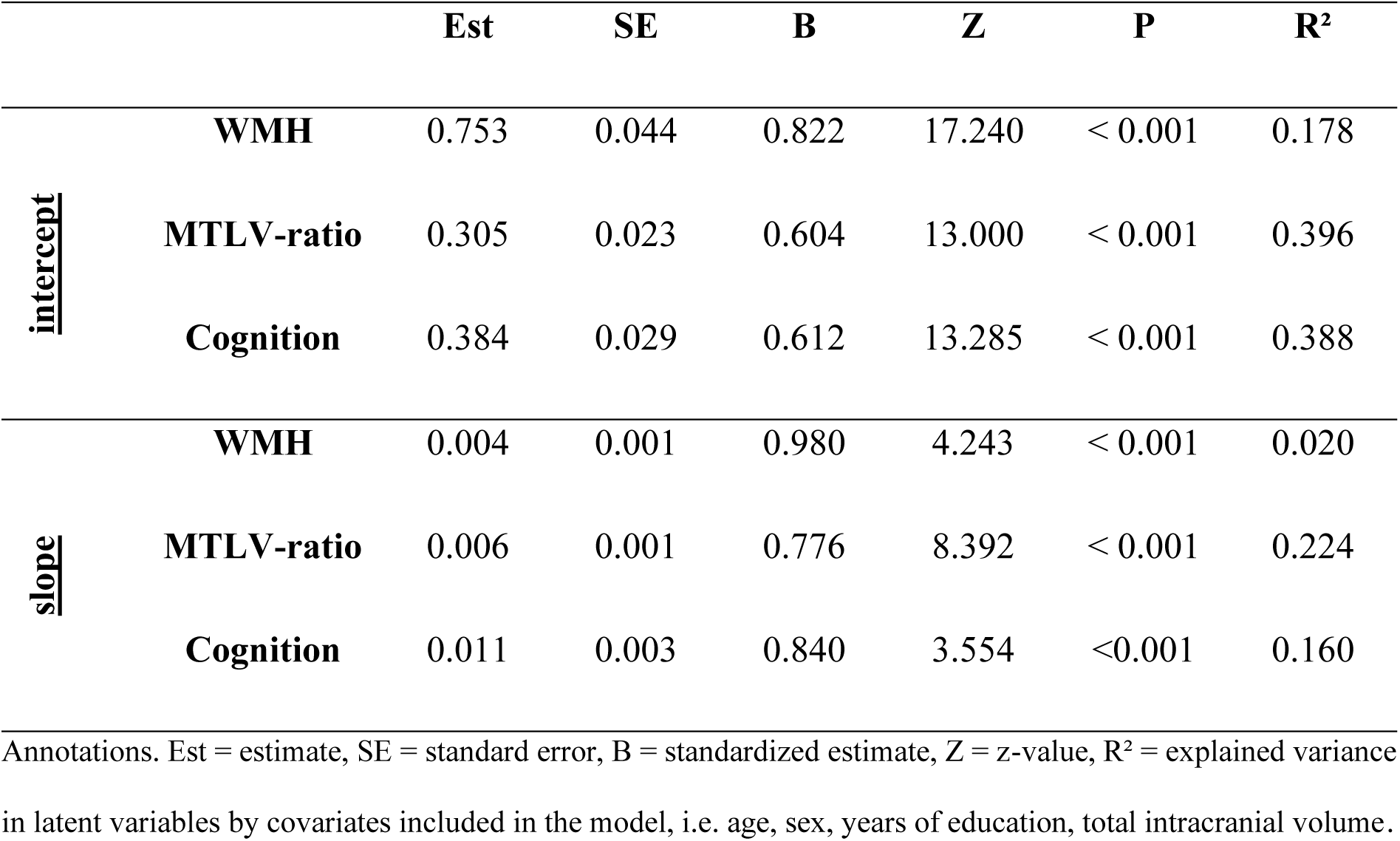
Variance of latent intercepts and slopes of WMH, MTLV-ratio and cognition as assessed by PACC5.

|  |  | **Est** | **SE** | **B** | **Z** | **P** | **R<sup>2</sup>** |
| --- | --- | --- | --- | --- | --- | --- | --- |
| **intercept** | **WMH** | 0.753 | 0.044 | 0.822 | 17.240 | < 0.001 | 0.178 |
|  | **MTLV-ratio** | 0.305 | 0.023 | 0.604 | 13.000 | < 0.001 | 0.396 |
|  | **Cognition** | 0.384 | 0.029 | 0.612 | 13.285 | < 0.001 | 0.388 |
| **slope** | **WMH** | 0.004 | 0.001 | 0.980 | 4.243 | < 0.001 | 0.020 |
|  | **MTLV-ratio** | 0.006 | 0.001 | 0.776 | 8.392 | < 0.001 | 0.224 |
|  | **Cognition** | 0.011 | 0.003 | 0.840 | 3.554 | <0.001 | 0.160 |

Annotations. Est = estimate, SE = standard error, B = standardized estimate, Z = z-value, R<sup>2</sup> = explained variance in latent variables by covariates included in the model, i.e. age, sex, years of education, total intracranial volume.

#### 3.2.2 Trajectories of neurocognitive domains are associated to age, sex, and years of education

##### Age

At baseline, older individuals had lower MTLV-ratios (*age → intercept MTLV-ratio*: *β* = - 0.500, *Z* = -12.818, *p* < 0.001), higher WMH volumes (*age → intercept WMH*: *β* = 0.371, *Z* = 9.248, *p* < 0.001) and performed worse in PACC5 (*age → intercept Cognition*: *β* = -0.398, *Z* = -9.620, *p* < 0.001). Over time, they tended to undergo faster declines in MTLV-ratios (*age → slope MTLV-ratio*: *β* = -0.388, *Z* = -7.832, *p* < 0.001), and show lower PACC5 performance changes (*age → slope Cognition*: *β* = -0.370, *Z* = -4.739, *p* < 0.001).

##### Female sex

Females yielded better PACC5 performance at baseline (*sex → intercept Cognition*: *β* = 0.356, *Z* = 9.011, *p* < 0.001), higher MTLV-ratio (*sex → intercept MTLV-ratio*: *β* = 0.145, *Z* = 3.065, *p* = 0.002), but also higher initial WMH volumes (*sex → intercept WMH*: *β* = 0.166, *Z* = 2.971, *p* = 0.003).

##### Education

Individuals with more years of education had better initial PACC5 performance (*years of education → intercept Cognition*: *β* = 0.257, *Z* = 6.343, *p* < 0.001), but did not show increased PACC5 performance changes (*years of education → slope Cognition*: *β* = 0.119, *Z* = 1.547, *p* = 0.112). Structurally, they also had higher initial MTLV-ratios (*years of education → intercept MTLV-ratio*: *β* = 0.098, *Z* = 2.568, *p* = 0.010). Education did not relate to the intercepts or slopes of WMH.

#### 3.2.3 Relationships among growth factors of WMH, ageing-related atrophy, and cognition

Individuals with higher baseline WMH volumes had lower baseline MTLV-ratios (*intercept WMH ∼ intercept MTLV-ratio*: *cov_Standardized_* = -0.139, *Z* = -3.261, *p* = 0.001) and experienced steeper declines in MTLV-ratios over time (*intercept WMH ∼ slope MTLV-ratio*: *cov_Standardized_* = -0.179, *Z* = -3.725, *p* < 0.001; **Figure 1A**). More emphasised rates of decline in MTLV-ratios were also observed in those with faster WMH progression (*slope WMH ∼ slope MTLV-ratio*: *cov_Standardized_* = -0.181, *Z* = -3.005, *p* = 0.003).

Individuals with better initial PACC5 performance showed higher baseline MTLV-ratios (*intercept cognition ∼ intercept MTLV-ratio*: *cov_Standardized_* = 0.153, *Z* = 2.908, *p* = 0.004) and less MTLV-ratio decline (*intercept cognition ∼ slope MTLV-ratio*: *cov_Standardized_* = 0.145, *Z* = 2.601, *p* = 0.009), the latter implying an ameliorating effect of cognitive capability on ageing-related atrophy.

PACC5 performance changes were more pronounced in individuals with higher MTLV-ratios at baseline, and with slower declines in MTLV ratios (*slope cognition ∼ intercept MTLV-ratio*: *cov_Standardized_* = 0.257, *Z* = 2.781, *p* = 0.005; *slope cognition ∼ slope MTLV-ratio*: *cov_Standardized_* = 0.483, *Z* = 4.207, *p* < 0.001; **Figure 1A & Table 2 & Figure 2**). Additionally slower progression of WMH was linked to higher PACC5 performance changes (*slope cognition ∼ slope WMH*: *cov_Standardized_* = -0.200, *Z* = -2.097, *p* = 0.036; **Figure 1A & Table 2 & Figure 2**). Collectively, these associations indicate that stronger increase of WMH and MTLV-ratio decline could both impede positive PACC5 performance changes.

**Table 2.** Independent effects of WMH and MTLV-ratio on cognitive changes in ageing. The table on the left shows the additive multiple linear regression of latent PACC5 slopes regressing on latent WMH and MTLV-ratio slopes, while controlling for latent intercepts of all neurocognitive domains. The figure on the right shows a 3D-Scatterplot of factor scores of slopes of WMH, MTLV-ratio and cognition. Colours reflect the slope in cognition with red colours pointing to negative and blue colours to positive slope estimates. Factor scores were extracted from the trivariate LGCM via regression-based method adjusted for effects of age, sex, years of education, and in the case of WMH and MTLV-ratio for total intracranial volume (TICV).

|  | **b** | **SE** | **T** | **p** |
| --- | --- | --- | --- | --- |
| **Intercept** | 0.130 | 0.008 | 15.495 | < 0.001 |
| **Slope WMH** | -0.267 | 0.062 | -4.334 | < 0.001 |
| **Slope MTLV-ratio** | 0.639 | 0.059 | 10.743 | < 0.001 |
| **Intercept PACC5** | 0.029 | 0.005 | 6.079 | < 0.001 |
| **Intercept WMH** | -0.003 | 0.004 | -0.803 | 0.422 |
| **Intercept MTLV-ratio** | -0.011 | 0.007 | -1.534 | 0.125 |

Annotations. b = regression coefficient, SE = standard error.

**Figure 2.**
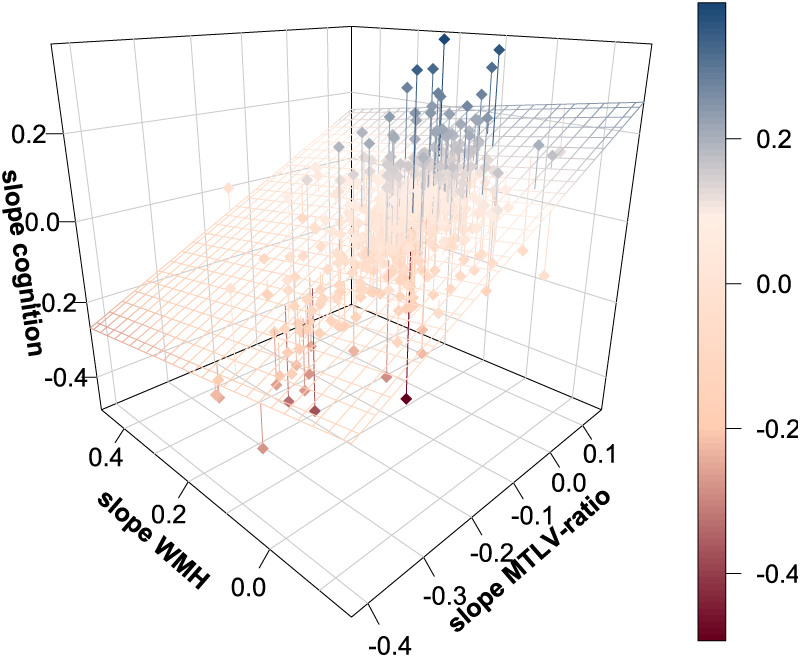
Independent effects of WMH and MTLV-ratio on cognitive changes in ageing. The table on the left shows the additive multiple linear regression of latent PACC5 slopes regressing on latent WMH and MTLV-ratio slopes, while controlling for latent intercepts of all neurocognitive domains. The figure on the right shows a 3D-Scatterplot of factor scores of slopes of WMH, MTLV-ratio and cognition. Colours reflect the slope in cognition with red colours pointing to negative and blue colours to positive slope estimates. Factor scores were extracted from the trivariate LGCM via regression-based method adjusted for effects of age, sex, years of education, and in the case of WMH and MTLV-ratio for total intracranial volume (TICV).

#### 3.2.4 Unique contributions of ageing-related atrophy and WMH to cognitive ageing

In order to assess the specific contributions of changes in WMH and MTLV-ratio to cognitive change, we conducted a multiple linear regression analysis, controlling for effects of latent intercepts (*F*(5, 499) = 70.95, *p* < 0.001, *R²_adjusted_* **=** 0.410; **Table 2**). It revealed that latent rates of changes of PACC5 were uniquely associated with each brain-level domain slope, i.e. MTLV-ratio (*b* = 0.639, *SD* = 0.059, *p* < 0.001) and WMH (*b* = -0.267, *SD* = 0.062, *p* < 0.001) (**Table 2 & Figure 2**). We did not observe indications for a detrimental interaction of these two processes (*F*(1, 498) = 1.372, *p* = 0.242; *ΔAIC* = 0.610).

### 3.3 Modifiable lifestyle factors and personality traits associate to changes in neurocognitive domains

After characterizing individual’s neurocognitive ageing trajectories in three domains and their interrelations, we examined which modifiable lifestyle factors and personality traits related to progression of WMH, MTLV-ratio decline or PACC5 changes specifically and generally.

First, we used the domain-specific extracted factor scores of latent slopes and correlated them with the lifestyle factors (**Figure 3**; correlation with latent intercepts and lifestyle factors in **Supplementary** Figure 4). We used partial spearman’s correlations to adjust for the effects of age, sex, years of education, and TICV (**Supplementary** Figure 5 shows full correlation matrix). We show unadjusted correlations in **Supplementary** Figure 6**&7**.

**Figure 3.**
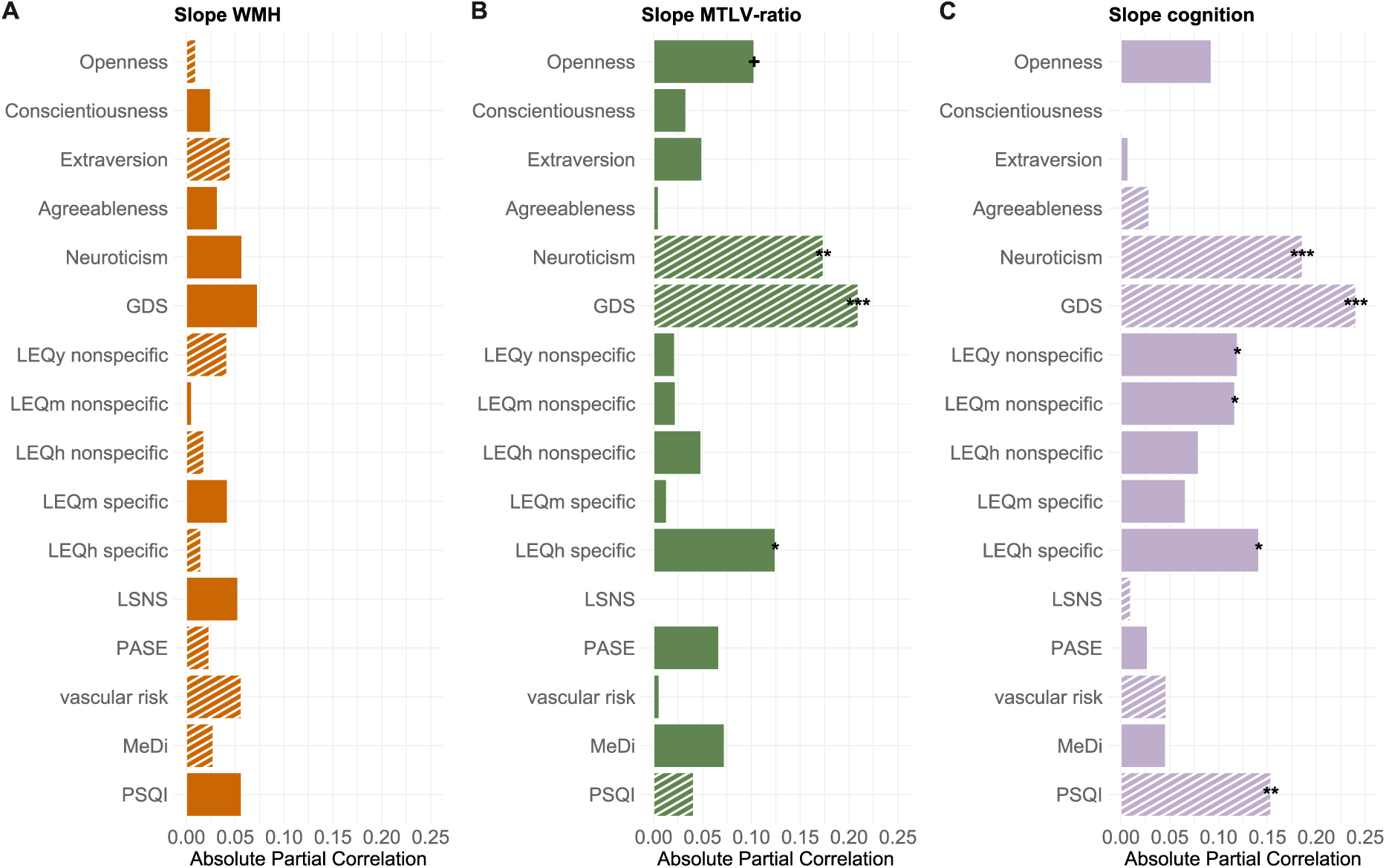
Associations of latent changes in neurocognitive domains with modifiable lifestyle factors. Factor scores for latent slopes were derived from the trivariate LGCM via regression-based method. We used partial Spearman’s correlations to account for the effects of age, sex, years of education, and total intracranial volume (TICV). All correlations were FDR-corrected. Filled bars denote positive correlation coefficients, striped bars denote negative correlation coefficients. Panels show relations between lifestyle factors and slopes of **(A)** total WMH, **(B)** MTLV-ratio, and **(C)** cognition as assessed with the PACC5. *** p < 0.001, ** p < 0.01, * p < 0.05, + p < 0.1. Personality traits were acquired via the Big Five Inventory BFI-10. GDS = geriatric depression scale. LEQ = lifetime experiences questionnaire assessed for three life periods y = young adulthood (13-30 years), m = midlife (30-65 years), h = late life (≥ 65 years or from retirement onward). LSNS = Lubben social network scale. PASE = physical activity scale for the elderly. MeDi = Mediterranean diet. PSQI = Pittsburgh sleep quality index (CAVE: by convention higher values denote lower sleep quality).

We did not observe any lifestyle factors to be associated with WMH progression after correction for multiple comparison. Yet, we found multiple modifiable lifestyle factors to be associated with changes in MTLV-ratio and PACC5. First, engaging in cognitively demanding leisure time activities in late life weakly related to lower MTLV-ratio decline (*ρ* = 0.125, *p_FDR_* = 0.024) and greater PACC5 performance changes (*ρ* = 0.141, *p_FDR_* = 0.009). Second, higher GDS scores and higher levels of neuroticism were linked to steeper MTLV-ratio decline (GDS: *ρ* = -0.210, *p_FDR_* < 0.001; neuroticism: *ρ* = -0.174, *p_FDR_* = 0.001) and lower PACC5 performance changes (GDS: *ρ* = -0.241, *p_FDR_* < 0.001; neuroticism: *ρ* = -0.186, *p_FDR_* < 0.001). Additionally, PACC5 performance changes appeared to weakly associate with leisure time activities in young adulthood (*ρ* = 0.119, *p_FDR_* = 0.032) and midlife (*ρ* = 0.117, *p_FDR_* = 0.035).

Leveraging multiple linear regression, we found that beyond age, sex, and years of education, the modifiable lifestyle factors of interest contributed low and non-significantly to variance in WMH changes (*F*(15,313) = 1.161, *p* = 0.302, *R²* = 0.053, *R²_adjusted_* = 0.007). Approximately 4.05% of variance in changes in MTLV-ratio decline were explained by modifiable lifestyle factors (*F*(15,313) = 1.922, *p* = 0.021, *R²* = 0.084, *R²_adjusted_* = 0.040). Lastly, modifiable lifestyle factors explained approximately 10.77% of variance in PACC5 changes (*F*(15,313) = 3.639, *p* < 0.001, *R²* = 0.1485, *R²_adjusted_* = 0.1077). **Supplementary** Figure 8 shows education-related differences in associations between lifestyle factors and personality traits and neurocognitive domains.

Next, we assessed the contributions of modifiable lifestyle factors and personality traits to individual differences in the brain-structure–related component of cognitive ageing, reflecting a domain-general brain maintenance index (**Figure 4**). Significant associations emerged for neuroticism (*ρ* = -0.167, *p_FDR_* < 0.001), depressive symptoms (*ρ* = -0.207, *p_FDR_* < 0.001), and engagement in cognitively demanding leisure time activities in late life (*ρ* = 0.133, *p_FDR_* = 0.017).

**Figure 4.**
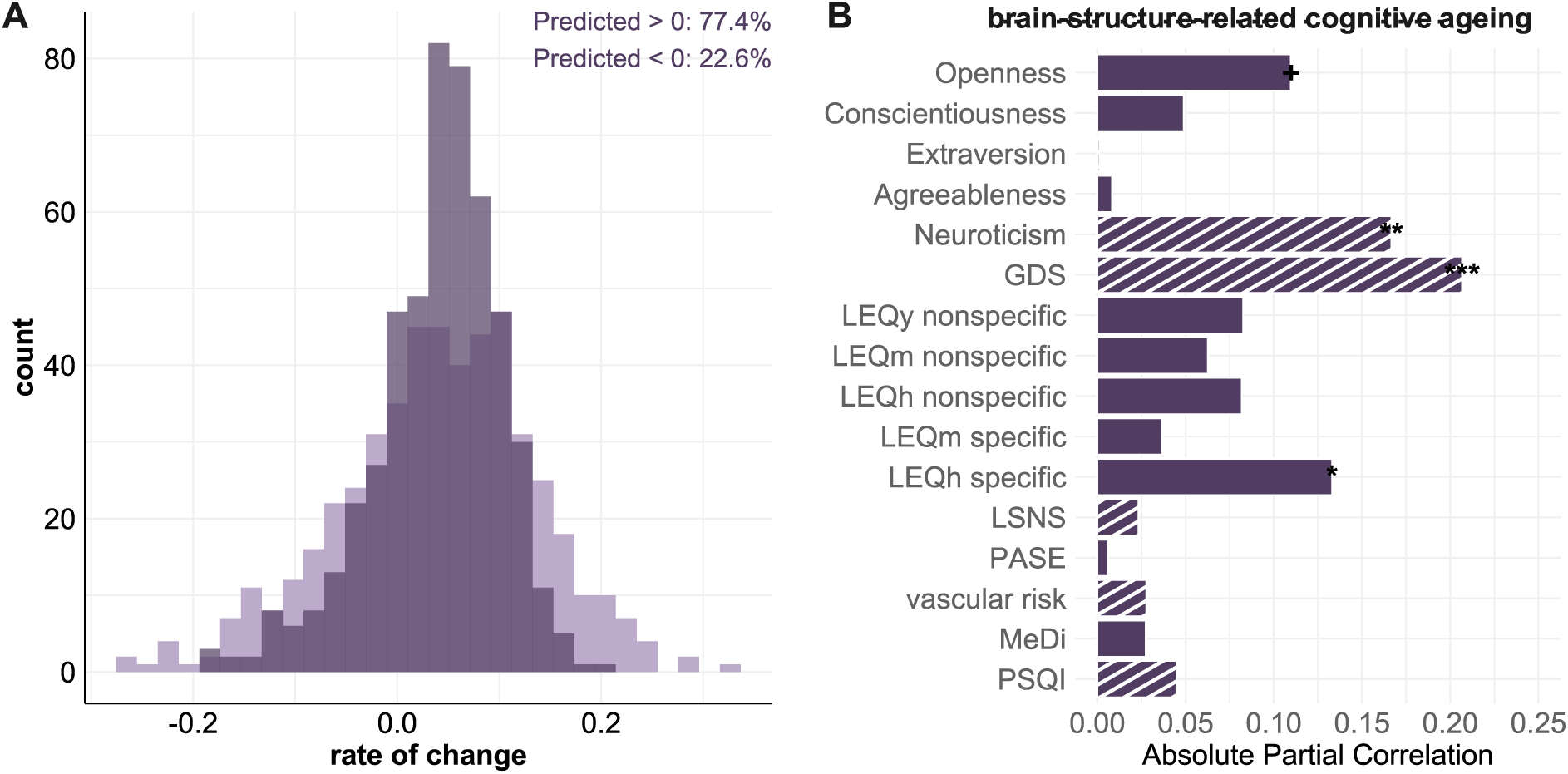
Individual maintenance as brain-structure–related cognitive ageing. Predicted values of cognitive slope were retrieved based on the multiple regression model, predicting cognitive slope by brain-domain changes and baseline levels of all neurocognitive domains (see 3.2.4), which reflect the brain-structure–related component of cognitive ageing. Higher individual predicted values hence indicate preserved cognitive functioning in proportion to preserved brain integrity, consistent with successful brain maintenance. **(A)** Distribution of the brain-structure–related component of cognitive ageing (darker purple) against factor scores of cognitive rate of change derived from the trivariate LGCM via regression-based method (lighter purple). **(B)** FDR-corrected partial Spearman’s correlations (accounting for the effects of age, sex, years of education, and total intracranial volume) of show relations between lifestyle factors and personality traits and brain-structure–related component of cognitive ageing. Filled bars denote positive correlation coefficients, striped bars denote negative correlation coefficients. *** p < 0.001, ** p < 0.01, * p < 0.05, + p < 0.1. Personality traits were acquired via the Big Five Inventory BFI-10. GDS = geriatric depression scale. LEQ = lifetime experiences questionnaire assessed for three life periods y = young adulthood (13-30 years), m = midlife (30-65 years), h = late life (≥ 65 years or from retirement onward). LSNS = Lubben social network scale. PASE = physical activity scale for the elderly. MeDi = Mediterranean diet. PSQI = Pittsburgh sleep quality index (CAVE: by convention higher values denote lower sleep quality).

## 4 Discussion

This study investigated brain maintenance by examining interrelated changes among WMH, ageing-related atrophy, and cognitive performance over four years in a cognitively unimpaired sample. We additionally identified modifiable lifestyle factors and personality traits associated with either domain-specific trajectories or a domain-general brain maintenance index, reflected in brain-structure–related cognitive ageing.

We observed significant WMH progression and declining MTLV-ratio over time, indicating ageing-related atrophy. These changes were interrelated, with WMH baseline levels notably contributing to ageing-related atrophy. Cognitive performance (PACC5) generally improved over time, however changes were limited by MTLV-ratio decline and WMH progression, highlighting their unique and joint impact on cognitive ageing, and relevance for brain maintenance ^16^. Neuroticism, depressive symptoms, and late-life cognitive engagement were key contributors to domain-specific trajectories and domain-general interindividual differences in brain maintenance.

Our findings underscore the interdependence of cerebrovascular and ageing-related atrophic changes in shaping cognitive trajectories, and suggest promising intervention targets to preserve brain health and cognitive function.

### 4.1 Role of demographics in neurocognitive ageing trajectories

Although WMH progressed in our cognitively unimpaired sample, interindividual variability in trajectories implied some individuals experienced greater progression ^13,49^, while others may show stable or regressing WMH ^50^. Female sex and higher age were related to increased WMH baseline burden, but not WMH progression ^51^. By assessing global WMH, regional WMH differences which relate to distinct risk profiles ^52,53^ might have been obscured.

MTLV-ratio decline was faster with increasing age, consistent with lifespan studies showing accelerated ventricular enlargement and MTL volume loss in older age ^1,5^.

Most individuals exhibited positive cognitive performance changes over time, though these diminished with higher age ^11,14^. Cognitive performance changes likely reflect a combination of both age-related decline and repetition-related practice effects. Notably, reduced practice effects have been linked to cognitive decline and an increased risk of dementia ^54,55^.

### 4.2 Relationship between ageing-related atrophy and WMH

WMH and MTLV-ratio levels and changes were significantly associated, even when controlling for shared risk factors ^28,30,56^. While such links have been described previously ^13,28,57,58^, our focus on global WMH precludes addressing potential regional associations with ageing-related atrophy. Such effects may arise from WMH intersecting distinct fibre tracts ^29,56^ or differences in their underlying aetiology ^6,52,53^.

Higher baseline levels of WMH were associated with steeper MTLV-ratio decline, reinforcing the assumption of that region’s vulnerability to cerebrovascular abnormalities ^24,30,59^. Our findings align with the notion that cerebrovascular changes may play an early and exacerbating role in ageing-related or neuropathological cascades ^58,60,61^, and might therefore serve as a proxy for brain maintenance ^16,61^.

We emphasise, however, that the relationship between WMH and ageing-related atrophy may vary by sample characteristics ^10,34,49,57^, and that latent pathology—e.g. in later AD stages— may overshadow the impact of early cerebrovascular abnormalities ^30^.

### 4.3 Associations of brain domain changes with cognitive changes

We presented evidence for brain maintenance, demonstrating that changes in MTLV-ratio and WMH were linked to and independently contributed cognitive performance changes ^16,31^. WMH progression—rather than baseline WMH ^13^—was linked to reduced cognitive performance changes, emphasising the relevance of monitoring and managing WMH progression for mitigating cognitive decline. Conversely, both baseline levels and decline of the MTLV-ratio were associated with cognitive performance changes, underlining the importance of MTL integrity in maintaining cognitive function in ageing ^12^.

Consistent with studies in healthy elderly ^49^ and cerebral amyloid angiopathy patients ^62^, MTLV-ratio decline explained more variance in cognitive outcomes than WMH. However, the association between baseline WMH and MTLV-ratio decline suggests that cerebrovascular abnormalities may contribute to downstream structural brain changes, reinforcing the idea that early cerebrovascular interventions could promote brain maintenance ^58,60,61^. Indeed, longitudinal studies ^29,57^ support indirect pathways linking WMH to cognitive decline via structural brain changes.

Exploring the consequences and underlying causes of interindividual variability in trajectories of WMH and ageing-related atrophy—and scenarios in which cognitive function remains stable despite structural brain changes—will ultimately inform our understanding of cognitive reserve and brain maintenance ^16^.

### 4.4 Domain-specific and domain-general contributions of lifestyle and personality

Several lifestyle factors showed domain-specific and domain-general associations with neurocognitive ageing, offering insight into potential mechanisms of brain maintenance ^16^. In line with the concept of differential preservation ^14^, the extent to which cognitive functions and structural brain integrity are maintained during ageing may depend on the interindividual expression of specific lifestyle factors. To clarify how different lifestyle characteristics relate to neurocognitive ageing, we examined their associations with domain-specific trajectories and with a domain-general brain maintenance index.

Most notably, late-life depressive symptoms were consistently associated with more negative trajectories across the two domain-specific trajectories of cognitive performance ^22,63^ and MTLV-ratio decline ^1,22,63^, and the domain-general brain maintenance index, even in the presence of mild symptoms ^63^. These findings align with the well-established role of depression as a major dementia risk factor ^17,22^. Neuroticism showed a similar pattern of domain-specific and domain-general associations. Given its relation to depressive symptoms ^64–66^—particularly under chronic stress ^64^—and its role as a risk factor for cognitive impairment and dementia conversion ^65,66^, our findings underscore the importance of addressing mental health and considering personality traits linked to mental health vulnerability to promote brain maintenance.

Furthermore, complex cognitive engagement during late life was related to better MTLV-ratio integrity, cognitive outcomes ^18,19^, and the domain-general brain maintenance index. These findings corroborate cross-sectional evidence that environmental enrichment bolsters functional integrity of memory networks related to preserved memory performance in ageing^67^.

We also observed more selective associations with regards to domain-specific contributions of modifiable lifestyle factors. In this way, cognitively stimulating leisure activity during young adulthood and midlife related to more favourable cognitive trajectories ^18,19,67^. Conversely, education only contributed to baseline levels of cognition, not changes ^48^. Consistent with preserved differentiation ^14^, education might hence contribute to interindividual differences in baseline cognitive functioning, not the rate of cognitive change. In contrast, education was positively associated with both baseline levels and changes of MTLV-ratio, indicating that educational attainment could promote domain-specific brain reserve and maintenance ^13,16,49^.

Additionally, cognitive performance changes were compromised by poor sleep quality ^68^, aligning with evidence on its role as an early marker and potential contributor to neuropathological changes ^69,70^. Compromised sleep quality may also reflect mental health issues, including depression ^71^, potentially mediating its impact on cognition ^72^.

Together, our results emphasise the importance of mental health, stress-coping across the lifespan, and engaging in a cognitively enriched lifestyle to promote brain maintenance and reduce ageing-related decline ^16,47,73^. Importantly, they suggest that these factors contribute not only to isolated domain-specific changes but also to a broader, domain-general maintenance mechanism. Further research is needed to clarify the underlying mechanisms of these effects.

### 4.5 Limitations and open questions

Several limitations warrant consideration. First, although LGCMs can determine the co-evolution of constructs, they cannot assess the delayed effect of change in one construct on change in another at a later time point. Other frameworks, such as latent change score models ^74^, could enable more nuanced insights into causal sequences.

Second, given the exclusion criteria, DELCODE participants had low vascular risk, which may have underestimated certain associations, particularly involving WMH. Similar studies in various cohorts are therefore needed to generalize findings.

Third, while our analysis was based on a specific operationalization of the three neurocognitive domains, future research should examine lesion load within specific hubs of cognitive networks ^59^, expand the cognitive domains studied, and clarify the differential relevance of various CSVD markers ^8,9,49,57^. Other pathological processes not considered here could also influence the observed interrelationships, such as genetics ^75^, inflammation ^60^, or change of AD biomarkers over time ^60^. Future research into the combined impact of these factors will elucidate mechanisms of brain maintenance.

Fourth, the assessment of modifiable lifestyle factors was non-exhaustive, relying mainly on self-report questionnaires prone to retrospective biases (e.g., LEQ; PASE), and some measures (e.g., cardiovascular risk score) may have lacked precision. Missing data (e.g., MeDi) may have reduced statistical power. These limitations may have reduced our ability to detect stronger effects. Additionally, the relatively short follow-up period may have been insufficient to capture long-term effects, particularly for midlife or earlier lifestyle factors related to brain reserve ^47^. Moreover, the role of modifiable lifestyle factors in cognitive reserve in this context remains unaddressed, as does whether their effects are directional or bidirectional ^17,19^. Future studies could benefit from more comprehensive and objective lifestyle measures and longitudinal investigations—ideally spanning the full lifespan—to elucidate the mechanisms underlying preserved differentiation and differential preservation ^14^, the coupled dynamics among brain structure, cognition, and lifestyle ^49^, and their directionality.

### 4.6 Conclusion

We showed that WMH can co-evolve with regional ageing-related atrophy and may accelerate its progression. Preventing cerebrovascular changes might therefore reduce vulnerability to ageing-related atrophy or pathology-related neurodegeneration—key drivers of cognitive decline and dementia. Together, WMH progression and ageing-related atrophy contribute to cognitive decline, underscoring their relevance for brain maintenance. Importantly, modifiable lifestyle factors influenced ageing-related atrophy and late-life cognition in domain-specific and domain-general ways, offering insight into potential mechanisms of brain maintenance. Our findings highlight the importance of managing cerebrovascular and mental health while fostering cognitive engagement and considering personality traits linked to mental health vulnerability to promote brain maintenance. This approach could not only mitigate the impact of ageing-related processes, but also lower the risk of distinct pathological changes and their synergistic interactions during preclinical dementia stages, potentially delaying the onset of overt clinical symptoms and functional impairments.

## 6 Acknowledgements

We would like to express our gratitude to all DELCODE participants. We also thank the Max Delbrück Centre for Molecular Medicine in the Helmholtz Association (MDC), Freie Universität Berlin Centre for Cognitive Neuroscience Berlin (CCNB), Bernstein Center für Computional Neuroscience Berlin, Universitätsmedizin Göttingen Core Facility MR-Research Göttingen, Institut für Klinische Radiologie Klinikum der Universität München, and Universitätsklinikum Tübingen MR-Forschungszentrum.

ED and GZ are funded by the Deutsche Forschungsgemeinschaft (DFG, German Research Foundation; Project ID 425899996).

## 9 Consent Statement

All participants gave written informed consent in accordance with the Declaration of Helsinki prior joining the study. DELCODE is retrospectively registered at the German Clinical Trials Register (DRKS00007966, 04/05/2015). Ethics committees of the medical faculties of all participating sites, Berlin (Charité, University Medicine), Bonn, Cologne, Göttingen, Magdeburg, Munich (Ludwig-Maximilians-University), Rostock, and Tübingen, gave ethical approval for this work. The ethics committee of the medical faculty of the University of Bonn led and coordinated the process.

## 10 Data availability statement

The data that support this study are not publicly available, but may be provided upon reasonable request.

## 11 Author contributions

DELCODE study design: ED, AS, FJ

Conceptualisation: IM, JB, GZ

Methodology: IM, JB, GZ, RK Software: IM, JB, GZ

Image processing: JB, RY

Formal analysis: IM Investigation: IM, GZ

Supervision: GZ, ED

Project administration: GZ

Funding acquisition: ED

Resources: GZ, ED

Writing – original draft preparation: IM, GZ, JB, RK

Writing – review and editing: all authors

All authors read and approved the final manuscript

