## Supplementary Material for "Linking latent trajectories of ageing-related atrophy, white matter hyperintensities, and cognitive ageing over four years: Insights into brain maintenance"

**Table 1. Overview of contributors to trajectories of each neurocognitive domain in terms of modifiable lifestyle factors and personality traits.**

| Possible brain maintenance factor | Operationalized by | Description/Details | Score |
|---|---|---|---|
| **Total cardiovascular risk score** | information of baseline medical records | • Assessed at baseline<br>• Equally weighted score according to former or current smoking, presence of obesity, hyperlipidaemia, hypertension, diabetes<br>• Each condition coded as present or absent<br>• sum of present risk factors was corrected for amount of available information | • 0.0 to 1.0<br>• Higher values reflect higher cardiovascular risk |
| **Late life depressive symptoms** | 15-item German version of geriatric depression scale (GDS) (Yesavage et al., 1982) | • Assessed annually<br>• 5 to 10 points correspond to mild to moderate depressive symptoms<br>• 11 to 15 points denote severe depressive symptoms | • 0.0 to 15.0<br>• Higher values reflect higher depressive symptoms |
| **Mediterranean diet** | European Prospective Investigation of Cancer semi-quantitative food frequency questionnaire (Nöthlings et al., 2007) | • Assessed at baseline<br>• Nine food groups (vegetables, legumes, fruits and nuts, dairy products, cereals, meat and poultry, fish, alcohol, ratio of monounsaturated fatty acids and saturated fat)<br>• adherence to Mediterranean diet according (Wesselman et al., 2021) | • 0.0 to 9.0<br>• higher values denote stronger adherence to Mediterranean diet |
| **Physical activity** | German version of physical activity scale for the elderly (PASE) (Märki, 2004; Washburn et al., 1993) | • assessed annually<br>• queried physical activity during the week prior each visit<br>• individuals report time spent sitting, by foot, with different activities of mild to severe intensities<br>• CAVE: modified scoring procedure that did not include the number of hours spent with an activity, as the reported hours by DELCODE participants proved to be implausible | • higher values denote higher physical activity |
| **Sleep quality** | Pittsburgh Sleep Quality Index (PSQI) (Buysse et al., 1989) | • assessed annually<br>• queried four weeks prior each visit<br>• total score used as proxy for sleep quality | • 0.0 to 21.0<br>• higher values reflect lower sleep quality |

| | | | |
|---|---|---|---|
| **Social network** | 6-item German version of Lubben Social Network Scale (LSNS) (Lubben et al., 2006) | • assessed annually<br>• queries amount of family members and friends, with whom participants were in regular contact or received support from | • 0.0 to 30.0<br>• scores below 12 indicate social isolation. |
| **Lifetime experiences** | Lifetime experiences questionnaire (LEQ) (Valenzuela & Sachdev, 2007) | • LEQ measures of young adulthood and midlife assessed at baseline<br>• LEQ for late life assessed at each visit<br>• queries complex cognitive engagement during three life periods: young adulthood (13-30 years), midlife (30-65 years) and late life (≥ 65 years or from retirement onward)<br>• differentiates between specific and non-specific activities assessed for each period<br> o Specific activities refer to life-period-specific activities (education, employment)<br> o non-specific activities refer to activities that can span all life periods (leisure time activities, e.g., reading, artistic endeavours) | • higher values reflect higher cognitive engagement |
| **Personality traits** | Big Five Inventory BFI-10 (Rammstedt & John, 2007) | • assessed at baseline<br>• assessed personality traits openness, conscientiousness, extraversion, agreeableness, and neuroticism according to the five-factor model<br>• each trait assessed with 2 items which were averaged for each dimension | • range per each personality trait from 1.0 – 5.0<br>• higher values reflect higher manifestation of trait |

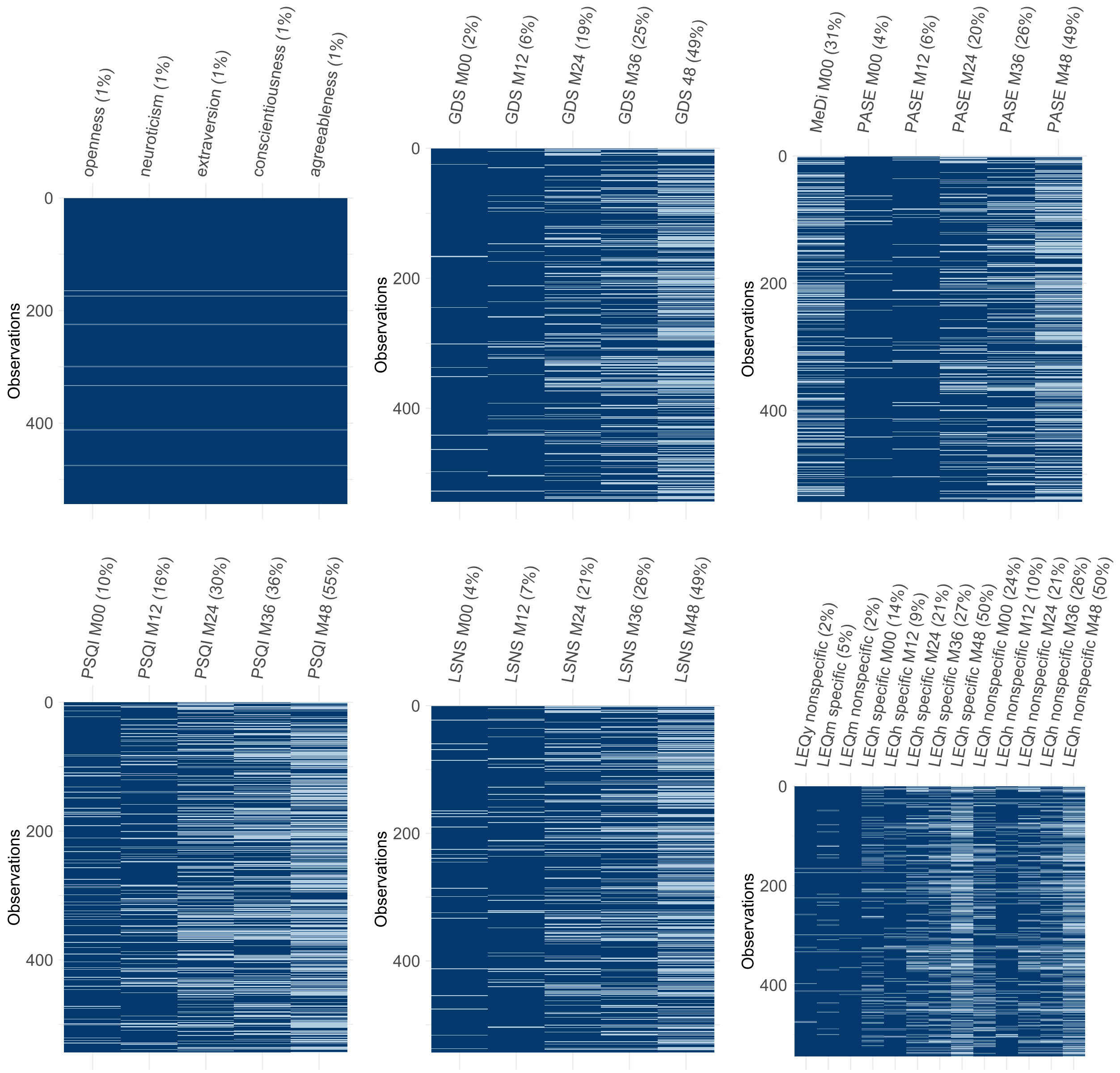


**Figure 1.** Missing data on modifiable lifestyle factors across measurement time points. M*n* denotes annual measurement time point (*n* = month of measurement/follow-up); M00 = Baseline. Personality traits were acquired via the BFI-10. GDS = geriatric depression scale. MeDi = Mediterranean diet. PASE = physical activity scale for the elderly. PSQI = Pittsburgh sleep quality index. LSNS = Lubben social network scale. LEQ = lifetime experiences questionnaire: y = young (13-30 years), m = middle-age (30-65 years), h = high age (≥ 65 years or from retirement onward).

**Table 2.** Descriptive statistics of personality traits and modifiable lifestyle factors in the total sample and comparing CU-stable and CU-converted individuals.

| | **Total** | **CU-stable** | **CU-converted** | $p_{\text{CU-stable vs. CU-converted}}$ |
|---|---|---|---|---|
| **cardiovascular risk** | **0.31 ± 0.22** | 0.29 ± 0.21 | 0.38 ± 0.24 | 0.002 |
| **Personality** | | | | |
| Openness | **3.57 ± 0.89** | 3.59 ± 0.89 | 3.42 ± 0.89 | 0.088 |
| Conscientiousness | **3.89 ± 0.80** | 3.88 ± 0.79 | 3.97 ± 0.85 | 0.293 |
| Extraversion | **3.25 ± 0.98** | 3.23 ± 0.98 | 3.29 ± 0.99 | 0.592 |
| Agreeableness | **3.25 ± 0.78** | 3.25 ± 0.78 | 3.31 ± 0.76 | 0.577 |
| Neuroticism | **2.98 ± 0.98** | 2.92 ± 0.99 | 3.18 ± 0.95 | 0.020 |
| **GDS** | **1.45 ± 1.77** | 1.25 ± 1.66 | 2.25 ± 2.07 | <0.001 |
| **MeDi** | **4.60 ± 1.58** | 4.65 ± 1.60 | 4.25 ± 1.61 | 0.111 |
| **PASE** | **31.37 ± 10.78** | 31.70 ± 10.80 | 30.03 ± 10.90 | 0.327 |
| **PSQI** | **5.57 ± 3.08** | 5.44 ± 2.97 | 5.82 ± 3.11 | 0.207 |
| **LSNS** | **21.87 ± 4.64** | 21.80 ± 4.61 | 22.1 ± 4.83 | 0.598 |
| **Lifetime experiences** | | | | |
| LEQy specific | **18.51 ± 5.31** | 18.60 ± 5.32 | 18.40 ± 5.38 | 0.608 |
| LEQm specific | **32.30 ± 10.86** | 32.60 ± 10.90 | 31.80 ± 10.90 | 0.592 |
| LEQh specific | **22.61 ± 4.69** | 22.80 ± 4.77 | 21.50 ± 4.46 | 0.927 |
| LEQy nonspecific | **18.91 ± 4.97** | 19.10 ± 5.09 | 18.10 ± 4.61 | 0.064 |
| LEQm nonspecific | **18.53 ± 4.70** | 18.60 ± 4.65 | 18.00 ± 5.12 | 0.196 |
| LEQh nonspecific | **15.94 ± 3.75** | 16.10 ± 3.80 | 15.20 ± 3.83 | 0.389 |

**Annotations.** Mean ± standard deviation. Comparison of CU-stable vs. CU-converted via Mann-Whitney-U tests on the averaged lifestyle factors. Personality traits were acquired via the BFI-10. GDS = geriatric depression scale. MeDi = Mediterranean diet. PASE = physical activity scale for the elderly. PSQI = Pittsburgh sleep quality index. LSNS = Lubben social network scale. LEQ = lifetime experiences questionnaire, y = young (13-30 years), m = middle-age (30-65 years), h = high age (≥ 65 years or from retirement onward).

**Table 3.** Model details of the main trivariate latent growth curve model with covariates age, sex, years of education, and total intracranial volume (TICV). Model fit: $\chi^2(151) = 213.56$, p = 0.001; CFI = 0.995; RMSEA = 0.028; SRMR = 0.018.

| | | | | Est | SE | B | Z | p |
|---|---|---|---|---|---|---|---|---|
| Associations with demographics | intercept WMH | ~ | Age | 0.355 | 0.038 | 0.371 | 9.248 | <0.001 |
| | | | Sex | 0.159 | 0.053 | 0.166 | 2.971 | 0.003 |
| | | | Years of education | -0.065 | 0.039 | -0.068 | -1.675 | 0.094 |
| | | | TICV | 0.248 | 0.051 | 0.260 | 4.882 | <0.001 |
| | slope WMH | ~ | Age | -0.006 | 0.004 | -0.101 | -1.446 | 0.148 |
| | | | Sex | 0.002 | 0.006 | -0.027 | 0.286 | 0.775 |
| | | | Years of education | -0.003 | 0.005 | -0.043 | -0.642 | 0.521 |
| | | | TICV | -0.003 | 0.005 | -0.049 | -0.631 | 0.528 |
| | intercept MTLV | ~ | Age | -0.355 | 0.028 | -0.500 | -12.818 | <0.001 |
| | | | Sex | 0.103 | 0.034 | 0.145 | 3.065 | 0.002 |
| | | | Years of education | 0.069 | 0.027 | 0.098 | 2.568 | 0.010 |
| | | | TICV | -0.136 | 0.031 | -0.191 | -4.320 | <0.001 |
| | slope MTLV | ~ | Age | -0.033 | 0.004 | -0.388 | -7.832 | <0.001 |
| | | | Sex | 0.006 | 0.005 | 0.065 | 1.073 | 0.283 |
| | | | Years of education | 0.007 | 0.004 | 0.077 | 1.485 | 0.137 |
| | | | TICV | -0.014 | 0.005 | -0.168 | -2.688 | 0.007 |
| | intercept cognition | ~ | Age | -0.315 | 0.033 | -0.398 | -9.620 | <0.001 |
| | | | Sex | 0.282 | 0.031 | 0.356 | 9.011 | <0.001 |
| | | | Years of education | 0.204 | 0.032 | 0.257 | 6.343 | <0.001 |
| | slope cognition | ~ | Age | -0.042 | 0.009 | -0.370 | -4.739 | <0.001 |
| | | | Sex | -0.008 | 0.009 | -0.070 | -0.916 | 0.360 |
| | | | Years of education | 0.013 | 0.009 | 0.119 | 1.547 | 0.122 |

| | | | | | | | | |
|---|---|---|---|---|---|---|---|---|
| **Associations latent variables** | intercept WMH | ~~ | slope WMH | -0.003 | 0.004 | -0.065 | -0.829 | 0.407 |
| | | | intercept MTLV | -0.066 | 0.020 | -0.139 | -3.261 | 0.001 |
| | | | intercept cognition | -0.034 | 0.023 | -0.062 | -1.431 | 0.152 |
| | | | slope MTLV | 0.012 | 0.003 | -0.179 | -3.725 | <0.001 |
| | | | slope cognition | -0.004 | 0.007 | -0.049 | -0.595 | 0.552 |
| | slope WMH | ~~ | intercept MTLV | -0.001 | 0.002 | -0.041 | -0.566 | 0.571 |
| | | | intercept cognition | 0.002 | 0.003 | 0.042 | 0.545 | 0.586 |
| | | | slope MTLV | -0.001 | 0.000 | -0.281 | -3.005 | 0.003 |
| | | | slope cognition | -0.001 | 0.001 | -0.200 | -2.097 | 0.036 |
| | intercept MTLV | ~~ | intercept cognition | 0.052 | 0.018 | 0.153 | 2.908 | 0.004 |
| | | | slope MTLV | 0.023 | 0.003 | 0.570 | 8.801 | <0.001 |
| | | | slope cognition | 0.015 | 0.005 | 0.257 | 2.781 | 0.005 |
| | intercept cognition | ~~ | slope MTLV | 0.007 | 0.003 | 0.145 | 2.601 | 0.009 |
| | | | slope cognition | 0.014 | 0.007 | 0.226 | 2.060 | 0.039 |
| | slope MTLV | ~~ | slope cognition | 0.004 | 0.001 | 0.483 | 4.027 | <0.001 |

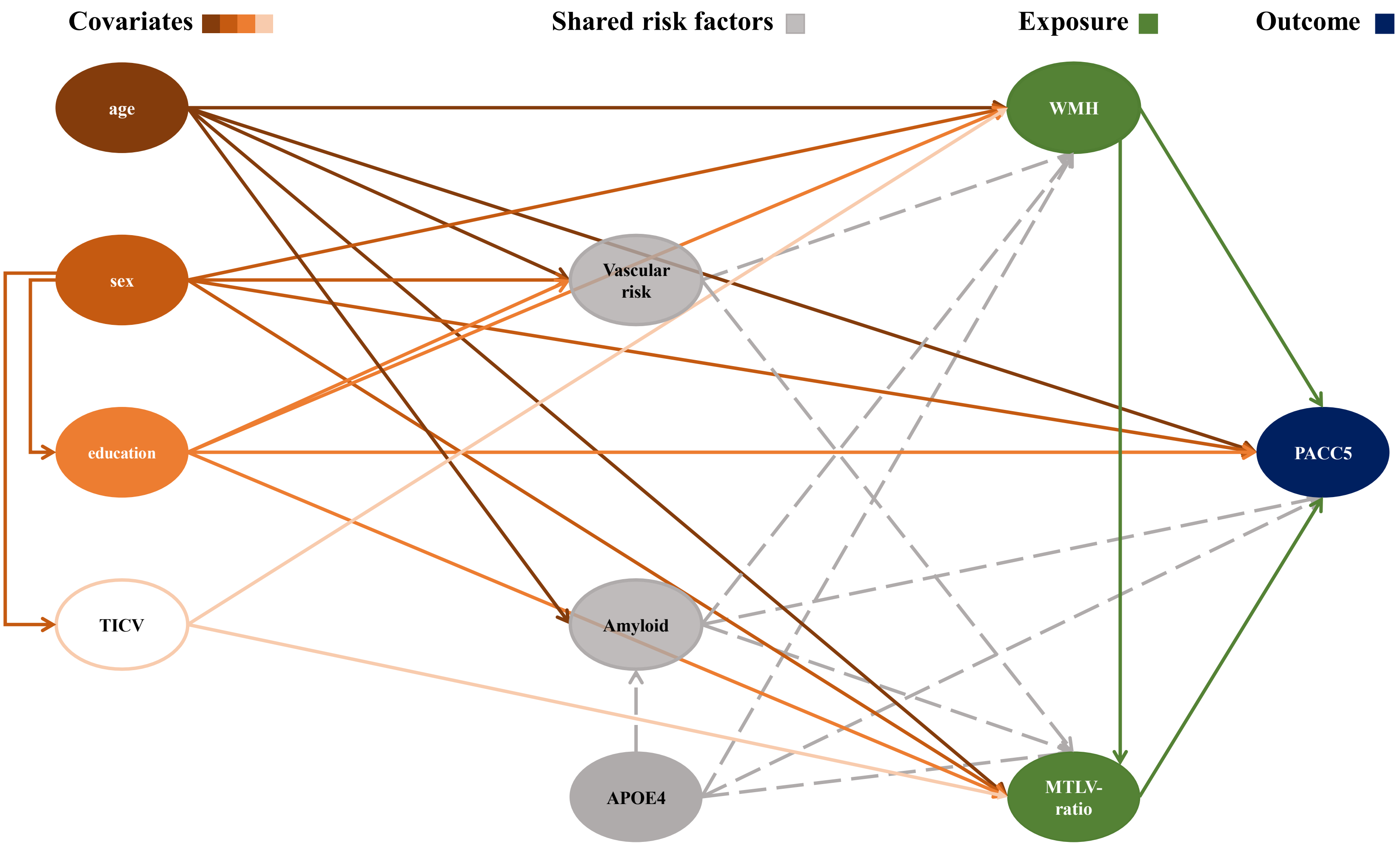


**Figure 2. Schematic directed acyclic graph (DAG).** Covariates were used in the main model. Inclusion of shared risk factors vascular risk, APOE4, and Plasma Amyloid was tested in subsequent additional models presented here in supplementary tables 3 and 4, respectively. TICV = total intracranial volume; WMH = white matter hyperintensities; MTLV-ratio = medial temporal lobe to ventricle ratio. PACC5 = preclinical Alzheimer's disease composite score.

**Table 4.** Model details of the trivariate latent growth curve model with additional covariate cardiovascular risk. Model fit: $\chi^2(164) = 225.66$, $p = 0.001$; CFI = 0.995; RMSEA = 0.026; SRMR = 0.019. Adding cardiovascular risk did not significantly improve model fit compared to the main model, $\Delta\chi^2(13) = 11.31$, $p = 0.585$.

| | | | | **Est** | **SE** | **B** | **Z** | **p** |
|---|---|---|---|---|---|---|---|---|
| **Associations with demographics** | intercept WMH | ~ | Age | 0.352 | 0.038 | 0.368 | 9.234 | <0.001 |
| | | | Sex | 0.174 | 0.054 | 0.181 | 3.227 | 0.001 |
| | | | Years of education | -0.053 | 0.039 | -0.056 | -1.364 | 0.172 |
| | | | Vascular risk | 0.075 | 0.038 | 0.078 | 1.970 | 0.049 |
| | | | TICV | 0.246 | 0.050 | 0.257 | 4.888 | <0.001 |
| | slope WMH | ~ | Age | -0.006 | 0.004 | -0.100 | -1.430 | 0.153 |
| | | | Sex | 0.001 | 0.006 | 0.022 | 0.228 | 0.820 |
| | | | Years of education | -0.003 | 0.004 | -0.047 | -0.702 | 0.482 |
| | | | Vascular risk | -0.002 | 0.004 | -0.025 | -0.372 | 0.710 |
| | | | TICV | -0.003 | 0.005 | -0.049 | -0.624 | 0.533 |
| | intercept MTLV | ~ | Age | -0.535 | 0.028 | -0.498 | -12.779 | <0.001 |
| | | | Sex | 0.097 | 0.034 | 0.137 | 2.856 | 0.004 |
| | | | Years of education | 0.064 | 0.028 | 0.090 | 2.319 | 0.020 |
| | | | Vascular risk | -0.032 | 0.025 | -0.045 | -1.244 | 0.214 |
| | | | TICV | -0.134 | 0.031 | -0.188 | -4.300 | <0.001 |
| | slope MTLV | ~ | Age | -0.033 | 0.004 | -0.388 | -7.873 | <0.001 |
| | | | Sex | 0.006 | 0.005 | 0.065 | 1.067 | 0.286 |
| | | | Years of education | 0.007 | 0.004 | 0.078 | 1.468 | 0.142 |
| | | | Vascular risk | -0.000 | 0.004 | -0.001 | -0.012 | 0.990 |
| | | | TICV | -0.014 | 0.005 | -0.167 | -2.674 | 0.007 |
| | intercept cognition | ~ | Age | -0.315 | 0.033 | -0.398 | -9.619 | <0.001 |
| | | | Sex | 0.282 | 0.031 | 0.356 | 9.012 | <0.001 |
| | | | Years of education | 0.204 | 0.032 | 0.258 | 6.345 | <0.001 |
| | slope cognition | ~ | Age | -0.042 | 0.009 | -0.371 | -4.741 | <0.001 |
| | | | Sex | -0.008 | 0.009 | -0.070 | -0.918 | 0.359 |
| | | | Years of education | 0.013 | 0.009 | 0.118 | 1.543 | 0.123 |

| | | | | | | | | |
|---|---|---|---|---|---|---|---|---|
| **Associations between latent variables** | intercept WMH | ~~ | slope WMH | -0.003 | 0.004 | -0.064 | -0.824 | 0.410 |
| | | | intercept MTLV | -0.063 | 0.020 | -0.133 | -3.138 | 0.002 |
| | | | intercept cognition | -0.031 | 0.023 | -0.057 | -1.307 | 0.191 |
| | | | slope MTLV | -0.012 | 0.003 | -0.179 | -3.684 | <0.001 |
| | | | slope cognition | -0.004 | 0.007 | -0.043 | -0.514 | 0.607 |
| | slope WMH | ~~ | intercept MTLV | -0.001 | 0.002 | -0.044 | -0.585 | 0.558 |
| | | | intercept cognition | 0.002 | 0.003 | 0.040 | 0.522 | 0.602 |
| | | | slope MTLV | -0.001 | 0.000 | -0.182 | -3.042 | 0.002 |
| | | | slope cognition | -0.001 | 0.001 | -0.202 | -2.121 | 0.034 |
| | intercept MTLV | ~~ | intercept cognition | 0.052 | 0.018 | 0.151 | 2.849 | 0.004 |
| | | | slope MTLV | 0.023 | 0.003 | 0.571 | 8.901 | <0.001 |
| | | | slope cognition | 0.014 | 0.005 | 0.253 | 2.738 | 0.006 |
| | intercept cognition | ~~ | slope MTLV | 0.007 | 0.003 | 0.147 | 2.582 | 0.010 |
| | | | slope cognition | 0.014 | 0.007 | 0.227 | 2.065 | 0.039 |
| | slope MTLV | ~~ | slope cognition | 0.004 | 0.001 | 0.483 | 4.030 | <0.001 |
| **Association covariates** | Age | ~~ | Sex | -0.194 | 0.042 | -0.194 | -4.030 | <0.001 |
| | | | Years of education | -0.111 | 0.046 | -0.111 | -2.432 | 0.015 |
| | | | Vascular risk | 0.098 | 0.044 | 0.098 | 2.212 | 0.027 |
| | | | TICV | 0.103 | 0.041 | 0.103 | 2.497 | 0.013 |
| | Sex | ~~ | Years of education | -0. 227 | 0.040 | -0.227 | -5.793 | <0.001 |
| | | | Vascular risk | -0.190 | 0.040 | -0.190 | -4.695 | <0.001 |
| | | | TICV | -0.665 | 0.039 | -0.665 | -33.182 | <0.001 |
| | Years of education | ~~ | Vascular risk | -0.112 | 0.041 | -0.112 | -2.736 | 0.006 |
| | | | TICV | 0.251 | 0.038 | 0.251 | 6.521 | <0.001 |
| | Vascular risk | ~~ | TICV | 0.123 | 0.045 | 0.123 | 2.749 | 0.006 |

Cardiovascular risk appeared to have a small effect on latent intercept of WMH, resulting in higher baseline levels of WMH with increased cardiovascular risk (vascular risk → intercept WMH: $\beta = 0.078$, $Z = 1.790$, $p = 0.049$). Adjusting for cardiovascular risk did not, however, result in changes to the qualitative interpretation of associations between latent intercepts and latent slopes between WMH, MTLV-ratio, and PACC5 performance (Aamand et al., 2024; Appelman et al., 2009; Bernal et al., 2024; Cox et al., 2019; Fiford et al., 2017; Godin et al., 2009).

**Table 5.** Model details of the trivariate latent growth curve model with additional covariates cardiovascular risk, APOE-ε4, and Plasma Aβ42/40. Model fit: $\chi^2(151) = 213.56$, p = 0.001; CFI = 0.995; RMSEA = 0.028; SRMR = 0.018. Adding cardiovascular risk, APOE-ε4, and Plasma Aβ42/40 did not significantly improve model fit compared to the main model, $\Delta\chi^2(35) = 21.71$, p = 0.962.

| | | | | **Est** | **SE** | **B** | **Z** | **p** |
|---|---|---|---|---|---|---|---|---|
| **Associations with demographics** | intercept WMH | ~ | Age | 0.340 | 0.039 | 0.356 | 8.730 | <0.001 |
| | | | Sex | 0.189 | 0.055 | 0.197 | 3.411 | 0.001 |
| | | | Years of education | -0.053 | 0.039 | -0.056 | -1.368 | 0.171 |
| | | | Vascular risk | 0.071 | 0.038 | 0.074 | 1.852 | 0.064 |
| | | | TICV | 0.241 | 0.050 | 0.252 | 4.795 | <0.001 |
| | | | APOE-ε4 | 0.017 | 0.041 | 0.017 | 0.405 | 0.685 |
| | | | Plasma Aβ42/40 | 0.065 | 0.049 | 0.068 | 1.338 | 0.181 |
| | slope WMH | ~ | Age | -0.006 | 0.004 | -0.096 | 1.328 | 0.184 |
| | | | Sex | 0.001 | 0.006 | 0.018 | 0.189 | 0.850 |
| | | | Years of education | -0.003 | 0.004 | -0.045 | -0.682 | 0.495 |
| | | | Vascular risk | -0.001 | 0.004 | -0.022 | -0.332 | 0.740 |
| | | | TICV | -0.003 | 0.005 | -0.051 | -0.641 | 0.522 |
| | | | APOE-ε4 | 0.003 | 0.005 | 0.043 | 0.587 | 0.557 |
| | | | Plasma Aβ42/40 | -0.001 | 0.005 | -0.023 | -0.288 | 0.773 |
| | intercept MTLV | ~ | Age | -0.356 | 0.028 | -0.502 | -12.821 | <0.001 |
| | | | Sex | 0.101 | 0.035 | 0.143 | 2.940 | 0.003 |
| | | | Years of education | 0.064 | 0.028 | 0.090 | 2.300 | 0.021 |
| | | | Vascular risk | -0.033 | 0.025 | -0.047 | -1.307 | 0.191 |
| | | | TICV | -0.131 | 0.031 | -0.185 | -4.195 | <0.001 |
| | | | APOE-ε4 | -0.035 | 0.027 | -0.050 | -1.335 | 0.182 |
| | | | Plasma Aβ42/40 | 0.020 | 0.032 | 0.029 | 0.643 | 0.520 |
| | slope MTLV | ~ | Age | -0.032 | 0.004 | -0.383 | -7.877 | <0.001 |
| | | | Sex | 0.004 | 0.005 | 0.048 | 0.791 | 0.429 |
| | | | Years of education | 0.006 | 0.004 | 0.072 | 1.395 | 0.163 |
| | | | Vascular risk | -0.000 | 0.004 | -0.005 | -0.124 | 0.901 |
| | | | TICV | -0.012 | 0.005 | -0.145 | -2.366 | 0.018 |
| | | | APOE-ε4 | -0.018 | 0.004 | -0.212 | -4.095 | <0.001 |
| | | | Plasma Aβ42/40 | -0.004 | 0.005 | -0.048 | -0.833 | 0.405 |

| | | | | | | | | |
|---|---|---|---|---|---|---|---|---|
| | intercept cognition | ~ | Age | -0.310 | 0.033 | -0.391 | -9.412 | <0.001 |
| | | | Sex | 0.276 | 0.033 | 0.349 | 8.316 | <0.001 |
| | | | Years of education | 0.204 | 0.032 | 0.257 | 6.337 | <0.001 |
| | | | APOE-ε4 | 0.009 | 0.032 | 0.011 | 0.282 | 0.778 |
| | | | Plasma Aβ42/40 | -0.026 | 0.038 | -0.033 | -0.683 | 0.494 |
| | slope cognition | ~ | Age | -0.040 | 0.009 | -0.351 | 4.444 | <0.001 |
| | | | Sex | -0.014 | 0.009 | -0.128 | -1.587 | 0.113 |
| | | | Years of education | 0.014 | 0.009 | 0.120 | 1.579 | 0.114 |
| | | | APOE-ε4 | -0.015 | 0.009 | -0.135 | -1.691 | 0.091 |
| | | | Plasma Aβ42/40 | -0.018 | 0.012 | -0.161 | -1.471 | 0.141 |
| **Associations between latent variables** | intercept WMH | ~~ | slope WMH | -0.003 | 0.004 | -0.064 | -0.839 | 0.402 |
| | | | intercept MTLV | -0.063 | 0.020 | -0.133 | -3.153 | 0.002 |
| | | | intercept cognition | -0.029 | 0.023 | -0.055 | -1.250 | 0.211 |
| | | | slope MTLV | -0.011 | 0.003 | -0.175 | -3.600 | <0.001 |
| | | | slope cognition | -0.002 | 0.007 | -0.028 | -0.322 | 0.748 |
| | slope WMH | ~~ | intercept MTLV | -0.001 | 0.002 | -0.040 | -0.546 | 0.585 |
| | | | intercept cognition | 0.002 | 0.003 | 0.040 | 0.526 | 0.599 |
| | | | slope MTLV | -0.001 | 0.000 | -0.179 | -2.893 | 0.004 |
| | | | slope cognition | -0.001 | 0.001 | -0.208 | -2.157 | 0.031 |
| | intercept MTLV | ~~ | intercept cognition | 0.051 | 0.018 | 0.150 | 2.861 | 0.004 |
| | | | slope MTLV | 0.023 | 0.003 | 0.579 | 8.998 | <0.001 |
| | | | slope cognition | 0.014 | 0.005 | 0.264 | 2.790 | 0.005 |
| | intercept cognition | ~~ | slope MTLV | 0.006 | 0.003 | 0.145 | 2.529 | 0.011 |
| | | | slope cognition | -0.029 | 0.023 | -0.055 | -1.250 | 0.211 |
| | slope MTLV | ~~ | slope cognition | 0.003 | 0.001 | 0.464 | 3.950 | <0.001 |

| | | | | | | | | |
|---|---|---|---|---|---|---|---|---|
| **Association between covariates** | Age | ~~ | Sex | -0.194 | 0.042 | -0.194 | -4.657 | <0.001 |
| | | | Years of education | -0.111 | 0.046 | -0.111 | -2.430 | 0.015 |
| | | | Vascular risk | 0.099 | 0.044 | 0.099 | 2.214 | 0.027 |
| | | | TICV | 0.103 | 0.041 | 0.103 | 2.500 | 0.012 |
| | | | APOE-ε4 | 0.009 | 0.043 | 0.009 | 0.206 | 0.837 |
| | | | Plasma Aβ42/40 | 0.240 | 0.049 | 0.240 | 4.863 | <0.001 |
| | Sex | ~~ | Years of education | -0.227 | 0.039 | -0.227 | -5.787 | <0.001 |
| | | | Vascular risk | -0.190 | 0.040 | -0.190 | -4.708 | <0.001 |
| | | | TICV | -0.665 | 0.020 | -0.665 | -33.203 | <0.001 |
| | | | APOE-ε4 | -0.071 | 0.043 | -0.071 | -1.638 | 0.101 |
| | | | Plasma Aβ42/40 | -0.313 | 0.040 | -0.313 | -7.823 | <0.001 |
| | Years of education | ~~ | Vascular risk | -0.112 | 0.041 | -0.112 | -2.736 | 0.006 |
| | | | TICV | 0.251 | 0.038 | 0.251 | 6.516 | <0.001 |
| | | | APOE-ε4 | 0.023 | 0.043 | 0.023 | 0.541 | 0.589 |
| | | | Plasma Aβ42/40 | 0.040 | 0.049 | 0.040 | 0.823 | 0.410 |
| | Vascular risk | ~~ | TICV | 0.123 | 0.045 | 0.123 | 2.757 | 0.006 |
| | | | APOE-ε4 | 0.012 | 0.044 | 0.012 | 0.272 | 0.786 |
| | | | Plasma Aβ42/40 | 0.136 | 0.049 | 0.136 | 2.756 | 0.006 |
| | TICV | ~~ | APOE-ε4 | 0.109 | 0.043 | 0.109 | 2.528 | 0.011 |
| | | | Plasma Aβ42/40 | 0.228 | 0.041 | 0.228 | 5.571 | <0.001 |
| | APOE-ε4 | ~~ | Plasma Aβ42/40 | 0.246 | 0.045 | 0.246 | 5.477 | <0.001 |

Plasma levels of Aβ42/40 were determined in a semi-automated two-step immunoprecipitation–immunoassay process encompassing the automated Aβ immunoprecipitation from plasma (CyBio FeliX liquid-handling instrument, Roboscreen, Leipzig, Germany) and the measurement of Aβ species via immunoassays (Mesoscale Discovery Aβ V-Plex immunoassay (6E10)). The procedure, its correlation with CSF-derived Aβ42/40 ratio, as well as its diagnostic and predictive utility in the DELCODE cohort are outlined in (Vogelgsang et al., 2024). Plasma levels of Aβ42/40 were available for 322 individuals. We opted for plasma instead of CSF-derived Aβ42/40, as the latter was only available in 185 individuals, and plasma Aβ42/40 has been shown to associate and discriminate well with regards to Aβ-PET (Doecke et al., 2020; Hu et al., 2022), especially when combined with APOE-ε4 status (Palmqvist et al., 2023; Vogelgsang et al., 2024; West et al., 2021). We reversed plasma Aβ42/40 levels to facilitate interpretation of effects, i.e., higher levels denote more pathological levels. We determined APOE-ε4 carriership based on the presence of at least one ε4 allele ($n_{carrier} = 150$; $n_{non\text{-}carrier} = 388$; $n_{missing} = 5$). The procedure for APOE genotyping is described in detail in (Jessen et al., 2018).

Adding cardiovascular risk, APOE-ε4, and Plasma Aβ42/40 simultaneously, resulted in the effect of cardiovascular risk on latent intercept of WMH becoming insignificant. Individuals with APOE-ε4 carriership had steeper declines in MTLV-ratios (APOE-ε4 → slope MTLV-ratio: $\beta = -0.212$, $Z = -4.095$, $p < 0.001$; (Cacciaglia et al., 2018; Gorbach et al., 2020; Schuff et al., 2008)). APOE-ε4 carriership was not associated with baseline levels of PACC5 performance, WMH, or MTLV ratio, nor with changes in PACC5 performance or WMH over time. There were no significant associations of more pathological baseline levels of plasma Aβ42/40 with baseline levels or changes in PACC5 performance, WMH, or MTLV-ratios. On the one hand, plasma Aβ42/40 is a valid but approximate measure CSF-Aβ42/40 levels (Vogelgsang et al., 2024) and hence might underrepresent AD pathological changes (Aschenbrenner et al., 2022). On the other hand, plasma Aβ42/40 might also relate to other risk constellations, e.g. cardiovascular risk (Li et al., 2024; Roher et al., 2009; Vogelgsang et al., 2024). In fact, we here found a relationship between plasma Aβ42/40 and our cardiovascular risk score (Vemuri et al., 2017; Wåhlin & Nyberg, 2019).We assume, that the effect might become significant, if AD biomarkers were derived from CSF (Aschenbrenner et al., 2022; Verberk et al., 2020). Taken together, adjusting for these variables simultaneously, did not, however, result in changes to the qualitative interpretation of associations between latent intercepts and latent slopes between WMH, MTLV-ratio, and PACC5 performance (Aamand et al., 2024; Appelman et al., 2009; Bernal et al., 2024; Cox et al., 2019; Fiford et al., 2017; Godin et al., 2009).

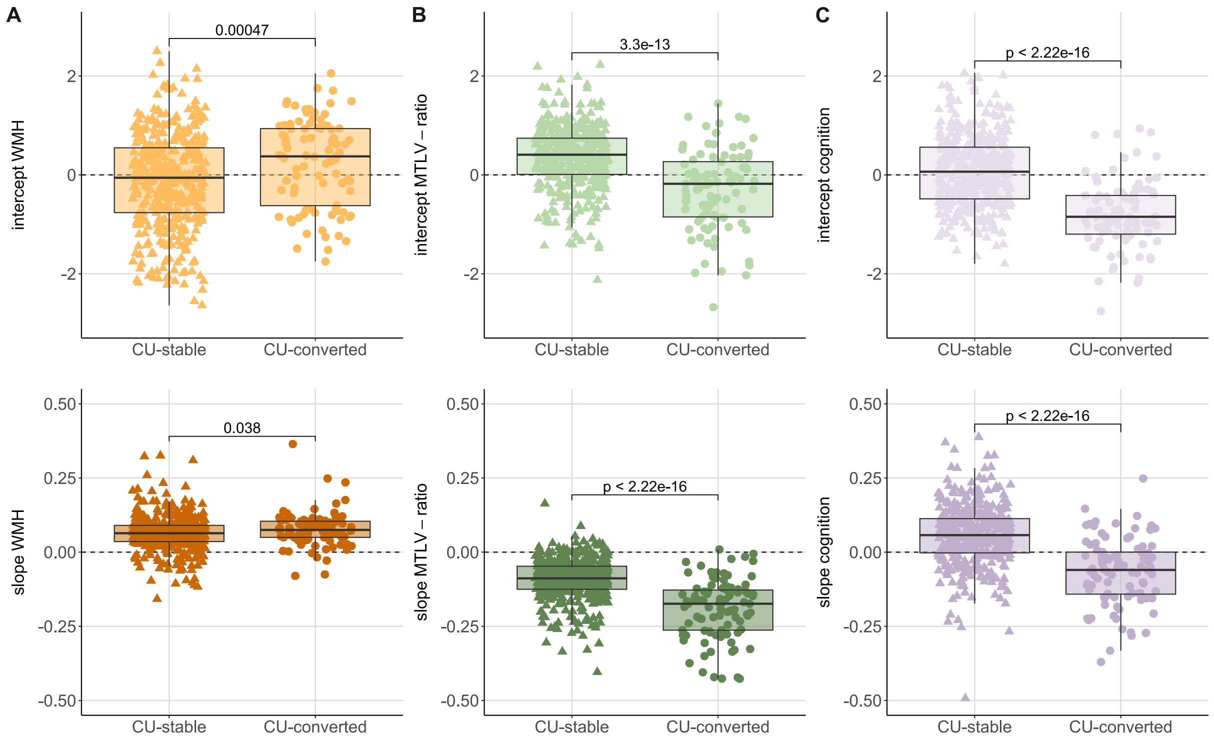


**Figure 3. Comparison of factor scores between cognitively stable and converting individuals reveal unfavourable pathological levels and progression within neurocognitive domains in individuals who converted to MCI or dementia during the course of the study as compared.**

Factor scores derived from the trivariate LGCM accounted for effects of age, sex, years of education, and in the case of WMH and MTLV-ratio for total intracranial volume (TICV). Due to the small sample of converted individuals, we refrained from using grouped LGCM to analyse these differences. Instead, we extracted regression-based factor scores of the latent intercepts and latent slopes and compared them via Mann-Whitney-U tests, with $\boldsymbol{r} = \frac{\boldsymbol{Z}}{\sqrt{\boldsymbol{N}}}$ as effect size. During the study, a total of 69 participants (22.52%) converted to MCI or dementia. Individuals who converted were generally older (U = 23836, p < 0.001), had higher cardiovascular risk (U = 23054, p = 0.002), and lower baseline plasma Aβ42/40 ratios (U = 8569, p < 0.001) than cognitively stable individuals. However, there were no differences between converting and cognitively stable individuals in terms of sex distribution ($\chi^2(1) = 1.24$, p = 0.266) or education (U = 17628, p = 0.213). Differences across groups in modifiable lifestyle factors are also detailed in **Supplementary Table 1**. **(A)** Total WMH differed between individuals who progressed to MCI or dementia and cognitively stable individuals regarding baseline levels ($U$ = 14751, $p$ < 0.001, $r$ = 0.155 [0.07 – 0.24]; (Bangen et al., 2018; Prosser et al., 2023)) and rates of changes ($U$ = 16564, $p$

= 0.038, $r = 0.092$ [0.01 – 0.18]; (Dadar et al., 2019; de Havenon et al., 2022; Jokinen et al., 2020; Kamal et al., 2023)). WMH progression was present in 94.62% of individuals converting to MCI or AD vs. in 89.35% of cognitively stable individuals (Fisher's Exact Test: non-significant). **(B)** MTLV-ratio differed between individuals who progressed to MCI or dementia and cognitively stable individuals regarding baseline levels ($U = 28479$, $p < 0.001$, $r = 0.324$ [0.24 – 0.40]) and rates of changes ($U = 30693$, $p < 0.001$, $r = 0.401$ [0.32 – 0.48]) (Bartos et al., 2019; Carmichael et al., 2007; Coupé et al., 2022; Driscoll et al., 2009; Macdonald et al., 2013; Mizuno et al., 2000; Schoemaker et al., 2019; Zahodne et al., 2015). MTLV-ratio decline was present in 98.92% of individuals converting to MCI or AD vs. in 92.22%. of cognitively stable individuals (Fisher's Exact Test: $OR = 6.68$, $p = 0.027$). **(C)** Cognition as assessed with the PACC5 differed between individuals who progressed to MCI or dementia and cognitively stable individuals regarding baseline levels ($U = 31462$, $p < 0.001$, $r = 0.428$ [0.36 – 0.50]) and rates of changes ($U = 30783$, p < 0.001, $r = 0.404$ [0.33 – 0.48]) (De Simone et al., 2021; Hassenstab et al., 2015; Jutten et al., 2020; Machulda et al., 2013; Samaroo et al., 2020). PACC5 performance gains were present in 25.81% of individuals converting to MCI or AD vs. in 74.82% of cognitively stable individuals (Fisher's Exact Test: $OR = 8.50$, p < 0.001).

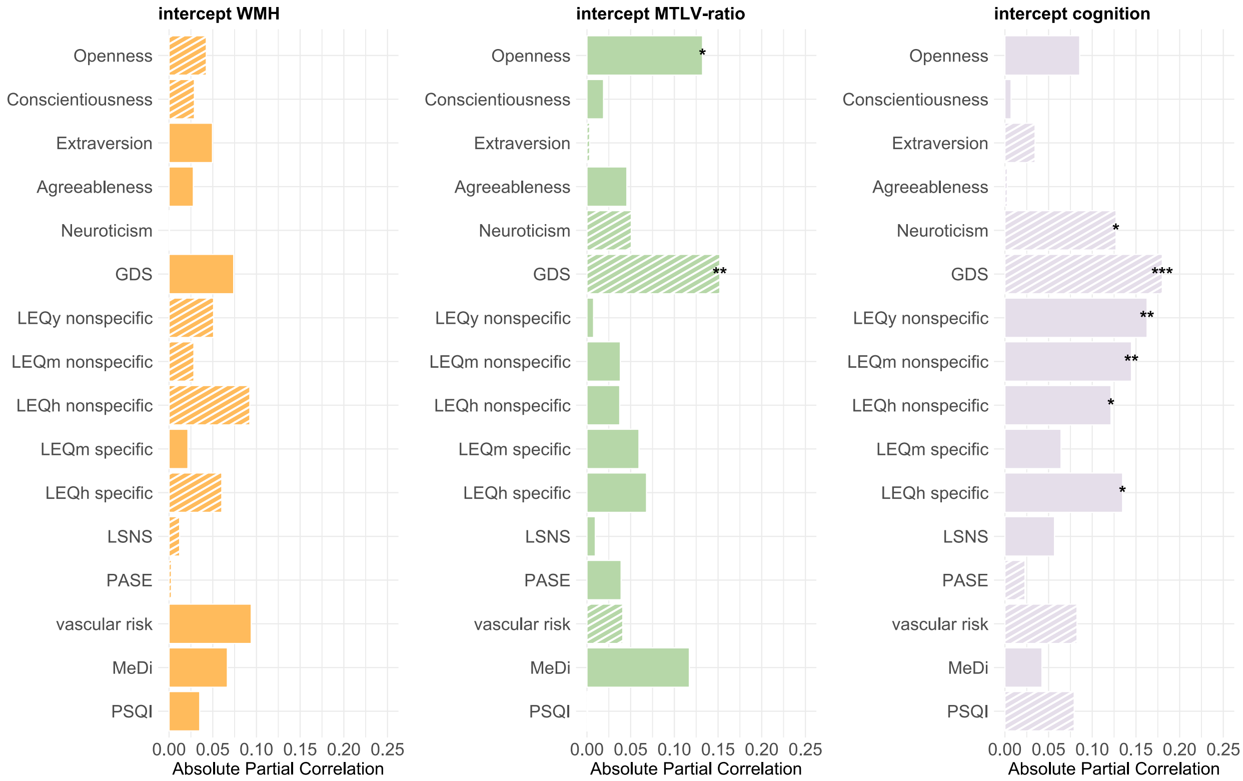


**Figure 4.** Associations of latent intercepts with modifiable lifestyle factors. Factor scores for latent intercepts were derived from the trivariate LGCM via regression-based method. We used partial Spearman's correlations to account for the effects of age, sex, years of education, and TICV. All correlations were FDR-corrected. Filled bars denote positive correlation coefficients, striped bars denote negative correlation coefficients. Panels show relations between lifestyle factors and intercepts of total WMH, MTLV-ratio, and cognition as assessed with the PACC5. *** $p < 0.001$, ** $p < 0.01$, * $p < 0.05$, + $p < 0.1$.

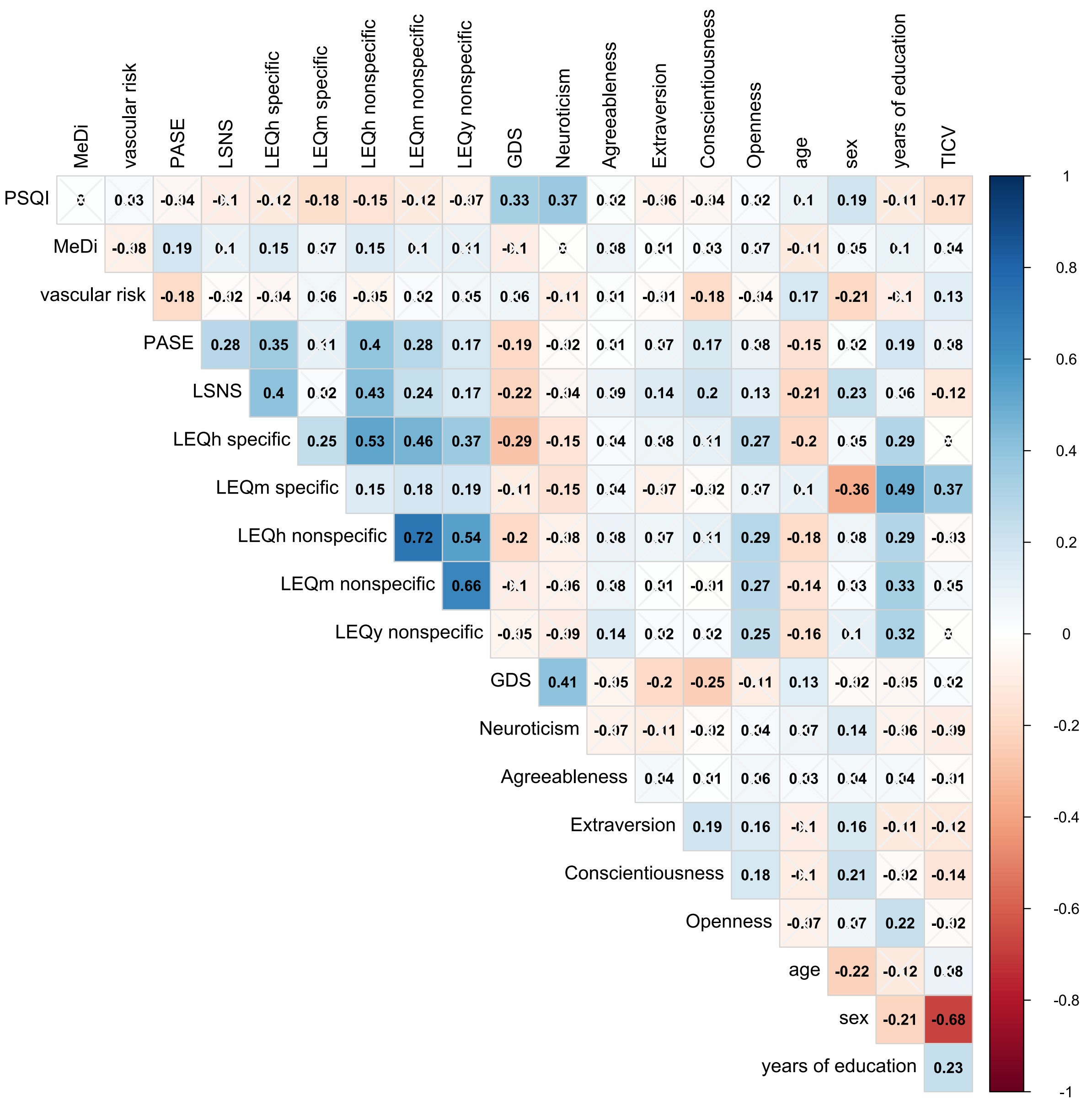

**Figure 5.** Correlation matrix of lifestyle factors and demographic factors. FDR-corrected Spearman correlations are shown. Non-significant correlations are crossed out.

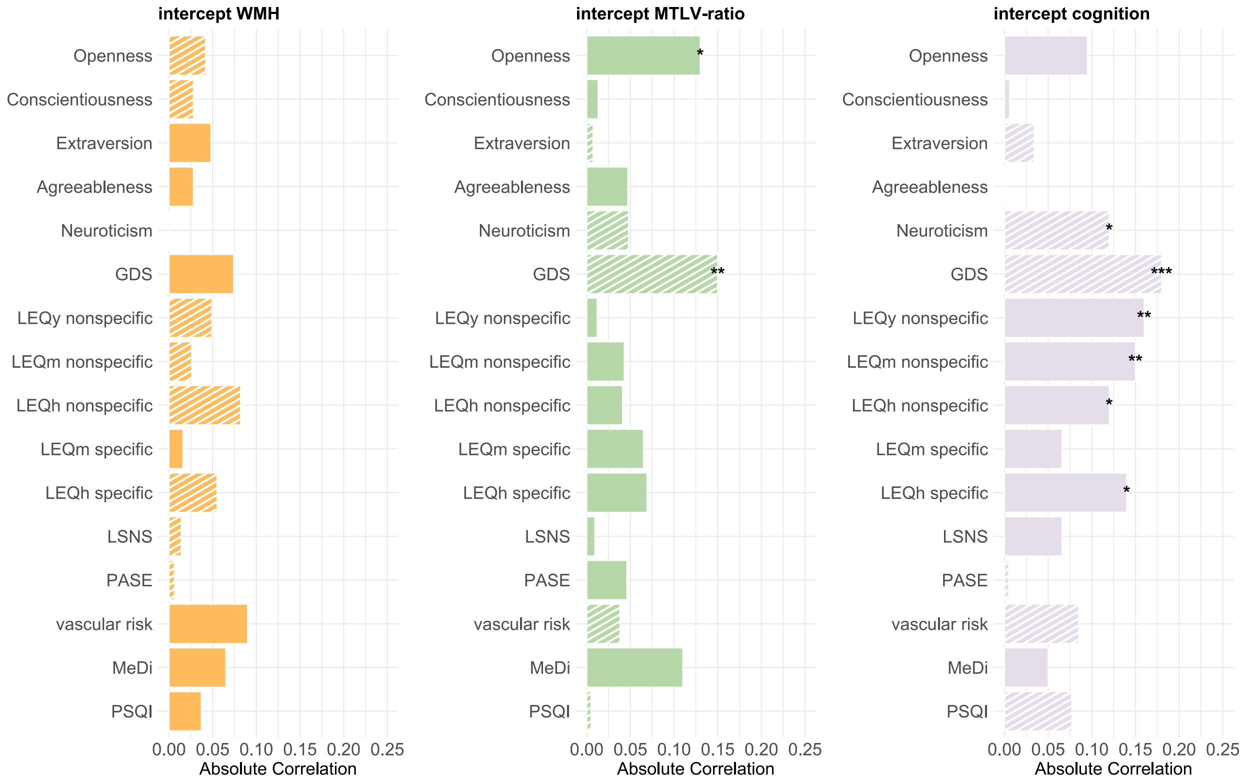


**Figure 6.** FDR-corrected Spearman's correlation of latent intercepts with modifiable lifestyle factors. Factor scores for latent intercepts were derived from the trivariate LGCM via regression-based method. Filled bars denote positive correlation coefficients, striped bars denote negative correlation coefficients. Panels show relations between lifestyle factors and intercepts of total WMH, MTLV-ratio, and cognition as assessed with the PACC5. *** $p < 0.001$, ** $p < 0.01$, * $p < 0.05$, + $p < 0.1$.

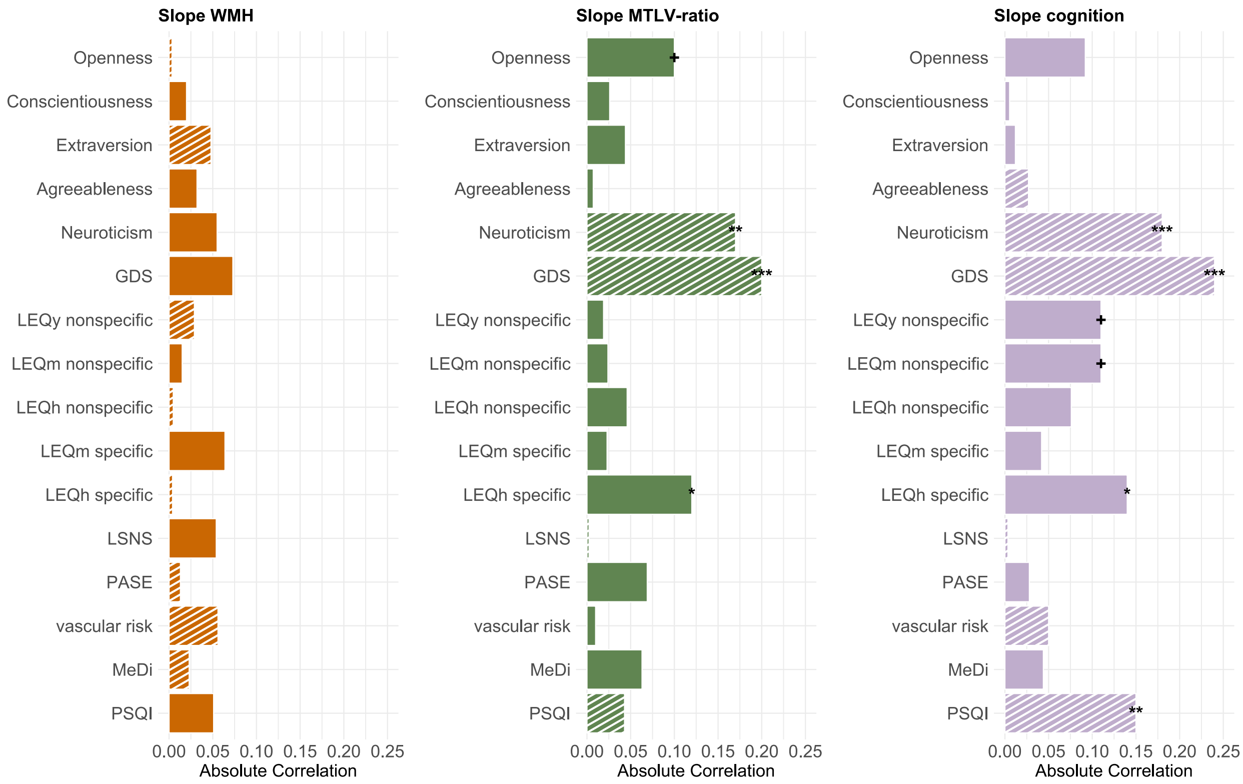


**Figure 7.** FDR-corrected Spearman's correlation of latent slopes with modifiable lifestyle factors. Factor scores for latent slopes were derived from the trivariate LGCM via regression-based method. Filled bars denote positive correlation coefficients, striped bars denote negative correlation coefficients. Panels show relations between lifestyle factors and latent slopes of total WMH, MTLV-ratio, and cognition as assessed with the PACC5. *** $p < 0.001$, ** $p < 0.01$, * $p < 0.05$, + $p < 0.1$.

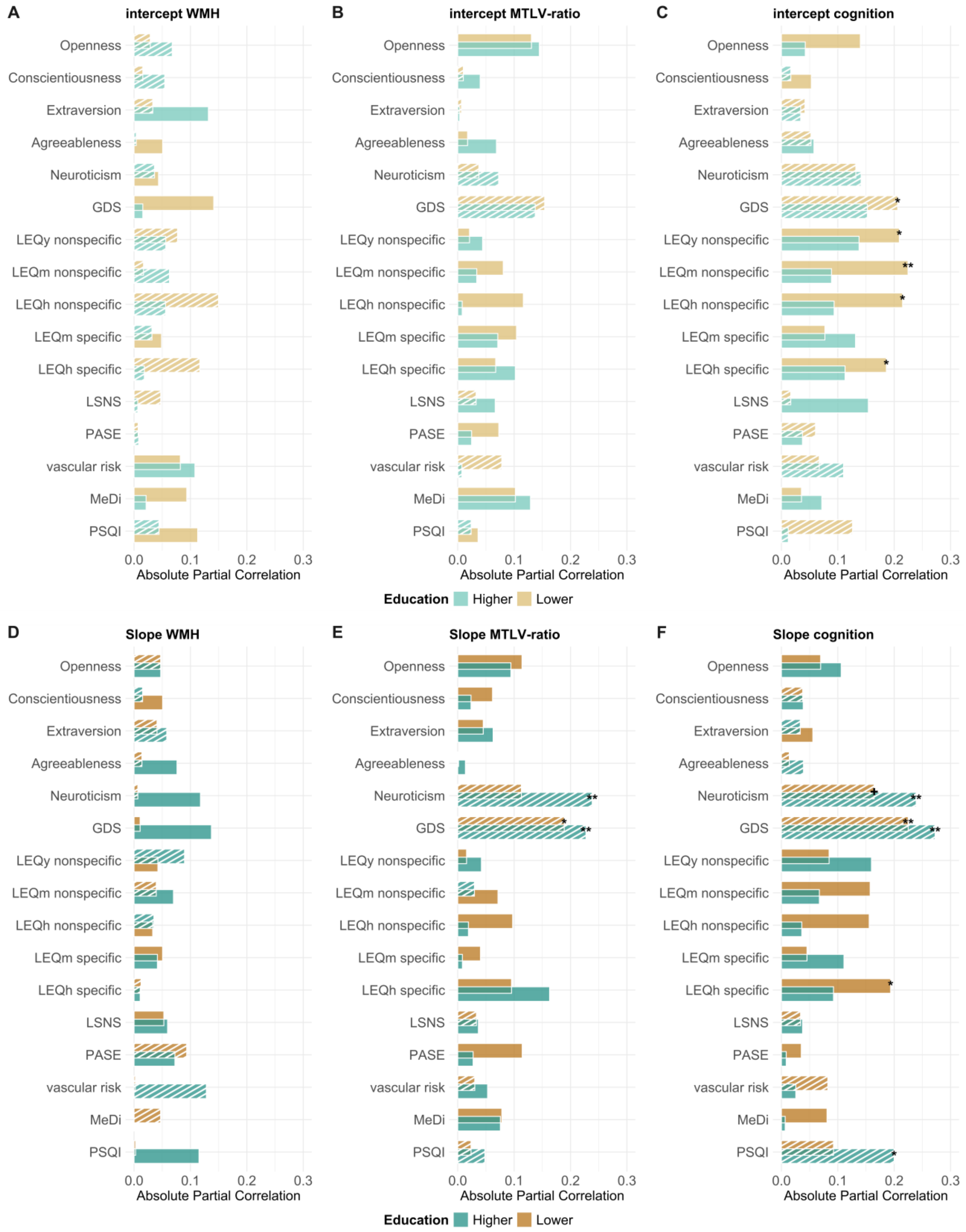


**Figure 8. Associations of neurocognitive domains with modifiable lifestyle factors across higher lower educated individuals.** Factor scores for latent intercepts and latent slopes were derived from a trivariate LGCM without correction for years of education via regression-based

method. We used grouped partial Spearman's correlations to account for the effects of age, sex, and total intracranial volume (TICV). Turquois bars represent correlations for higher education (Ø 17.4 years), amber bars represent correlations for lower education (Ø 12.3 years). All correlations were FDR-corrected. Filled bars denote positive correlation coefficients, striped bars denote negative correlation coefficients. Panels show relations between lifestyle factors, intercepts and slopes, respectively, of **(A&D)** total WMH, **(B&E)** MTLV-ratio, **(C&F)** cognition as assessed with the PACC5. *** $p < 0.001$, ** $p < 0.01$, * $p < 0.05$, + $p < 0.1$. Personality traits were acquired via the Big Five Inventory BFI-10. GDS = geriatric depression scale. LEQ = lifetime experiences questionnaire assessed for three life periods y = young adulthood (13-30 years), m = midlife (30-65 years), h = late life (≥ 65 years or from retirement onward). LSNS = Lubben social network scale. PASE = physical activity scale for the elderly. MeDi = Mediterranean diet. PSQI = Pittsburgh sleep quality index (CAVE: by convention higher values denote lower sleep quality). We were particularly interested in education-associated changes within neurocognitive domains. Neither in lower nor higher educated individuals, WMH progression was linked to modifiable lifestyle factors. In line with previous studies (Santos et al., 2014), we found that irrespective of level of education, higher GDS scores were related to steeper MTLV-ratio decline (lower: $\rho$ = -0.191, $p_{FDR}$ = 0.031; higher $\rho$ = -0.228, $p_{FDR}$ = 0.008) and lower PACC5 performance gains (lower: $\rho$ = -0.225, $p_{FDR}$ = 0.008; higher $\rho$ = -0.276, $p_{FDR}$ = 0.001). Higher levels of cognitively demanding leisure time activities in late life appeared to contribute to increased PACC5 performance changes in lower educated individuals ($\rho$ = 0.196, $p_{FDR}$ = 0.028), suggesting that engaging in cognitively stimulating behaviours could ameliorate cognitive decline in this group (Feng et al., 2024; Wang et al., 2024). In individuals with higher levels of education, neuroticism was associated with MTLV-ratio decline ($\rho$ = -0.238, $p_{FDR}$ = 0.007) and lower PACC5 performance changes ($\rho$ = -0.236, $p_{FDR}$ = 0.007). Moreover, increased PACC5 performance changes were also linked to better sleep quality in this group ($\rho$ = -0.209, $p_{FDR}$ = 0.018), as was reported in a previous study (Joo et al., 2022).

Feng, Y., Jia, S., Zhao, W., Wu, X., Zuo, Y., Wang, S., Zhao, L., Ma, M., Guo, X., Tarimo, C. S., Miao, Y., & Wu, J. (2024). Independent and Joint Associations of Socioeconomic Status and Lifestyle behaviors with Cognitive Impairment among Elderly Chinese Population. *Journal of Prevention of Alzheimer's Disease*, *5*(11), 1513–1522. https://doi.org/10.14283/jpad.2024.127

Joo, H. J., Kwon, K. A., Shin, J., Park, S., & Jang, S. I. (2022). Association between sleep quality and depressive symptoms. *Journal of Affective Disorders*, *310*(May), 258–265. https://doi.org/10.1016/j.jad.2022.05.004

Santos, N. C., Costa, P. S., Cunha, P., Portugal-Nunes, C., Amorim, L., Cotter, J., Cerqueira, J. J.,

Palha, J. A., & Sousa, N. (2014). Clinical, physical and lifestyle variables and relationship with cognition and mood in aging: a cross-sectional analysis of distinct educational groups. *Frontiers in Aging Neuroscience*, *6*(FEB), 1–15. https://doi.org/10.3389/fnagi.2014.00021

Wang, K., Fang, Y., Zheng, R., Zhao, X., Wang, S., Lu, J., Wang, W., Ning, G., Xu, Y., & Bi, Y. (2024). Associations of socioeconomic status and healthy lifestyle with incident dementia and cognitive decline: two prospective cohort studies. *EClinicalMedicine*, *76*, 102831. https://doi.org/10.1016/j.eclinm.2024.102831
